# Epigenetic misprogramming of cell differentiation links transient viral infection to chronic diseases across tissues

**DOI:** 10.64898/2026.09.08.26362522

**Authors:** Bernard Friedenson

**Affiliations:** Dept Biochemistry and Molecular Genetics and Illinois Cancer Center, College of Medicine University of Illinois Chicago, Chicago, Illinois

## Abstract

Virus-induced epigenetic changes in stem-cell differentiation pathways emerge here as a shared mechanism linking fundamentally different chronic diseases, offering candidate biomarkers and new intervention targets. Chronic diseases increasingly trace back to transient viral exposure, yet unrelated pathologies also arise in tissues showing no signs of infection. Using Epstein–Barr virus (EBV), I show that different chronic EBV-associated diseases share epigenetic misprogramming within stem-cell differentiation hierarchies. These alterations propagate across diverse cell lineages, creating latent vulnerabilities unmasked by secondary insults. Breast cancer, long debated for possible EBV involvement, carries EBV-like methylation signatures in both precursor and invasive lesions, independent of lymphocytic infiltration. In multiple sclerosis, three independent cohorts (non-pathologic brain tissue, normal-appearing white matter, and blood) showed methylation abnormalities in the same stem-cell differentiation pathways disrupted in proven EBV-driven tumors and breast cancer. These findings identify virus-induced lineage misprogramming as an occult cross-disease mechanism supported by direct primary evidence.

## Introduction

Viral infections are increasingly linked to chronic diseases, yet the mechanisms connecting transient infection to pathology in tissues that show no signs of infection remain unclear^1–3^. A prevalent view is that acute viral infections leave long-lasting epigenetic and transcriptional imprints on hematopoietic and immune cells that reshape future inflammatory responses^4–7^. Other existing explanations add complex sets of abnormalities in diverse uninfected tissues^8–10^.

Here, a more general mechanism is proposed: infections install durable epigenetic alterations in stem and progenitor cell differentiation pathways. Viruses exploit epigenetic vulnerabilities to rewire differentiation within stem-cell hierarchies regardless of inflammation and ongoing infection. Such disruptions misprogram lineage commitment, generating disease vulnerabilities that persist long after viral clearance, propagating through successive cell generations in diverse tissues. These changes link very different diseases, which then depend on secondary insults.

Epstein–Barr virus (EBV) provides the mechanistic model. EBV establishes lifelong latent infection in almost all humans, causes massive changes in host gene expression^11^, and contributes to cancers, neurologic disease, and autoimmunity. Latency occurs in distinct anatomical reservoirs, enabling disconnected diseases^12^ . Although EBV’s roles in nasopharyngeal carcinoma (NPC)^13–15^, endemic Burkitt lymphoma (BL)^16^ , and multiple sclerosis (MS)^17,18^ are well supported, these diseases have been studied in isolation and the mechanisms do not converge.

Here, I show that epigenetic misprogramming of differentiation pathways is a unifying mechanism across diseases that have never been considered mechanistically related. Breast cancer offers a critical test case because EBV involvement has been debated for decades with no mechanistic framework able to reconcile contradictory evidence^19–21^ or to explain breast metastasis from NPC^22^ . Yet breast cancer regulatory elements show striking EBV-like methylation patterns, emerging as early as the ductal carcinoma in situ (DCIS) precursor stage, long before malignant progression. The same epigenetic strategy extends to the CNS in MS, permeating non-pathologic brain tissue, normal appearing white matter (NAWM), and MS-derived DNA circulating in blood. Even cured, non-malignant keratinocytes retain EBV-like methylation signatures, revealing them as an intrinsic consequence of infection.

Together, these findings suggest that virus induced epigenetic reprogramming is a conserved upstream process that becomes imprinted on stem-cell hierarchy and shapes tissue specific disease susceptibility to secondary insults. The evidence presented below shows how consistently the mechanism occurs across fundamentally different diseases.

## Results

### Breast cancers share highly specific abnormal methylation sites with EBV-associated malignancies

Across autosomes, breast cancer gene control elements^23^ exhibited widespread alignment of abnormal methylation with differential methylation in NPC and BL. Many sites agreed within 10 bp and frequently had identical genomic coordinates, far exceeding chance expectations (Fig. 1a). Distribution level comparisons confirmed this concordance (p<0.0001), whereas randomized controls produced minimal or absent overlap (Fig. 1a).

**Fig. 1.**
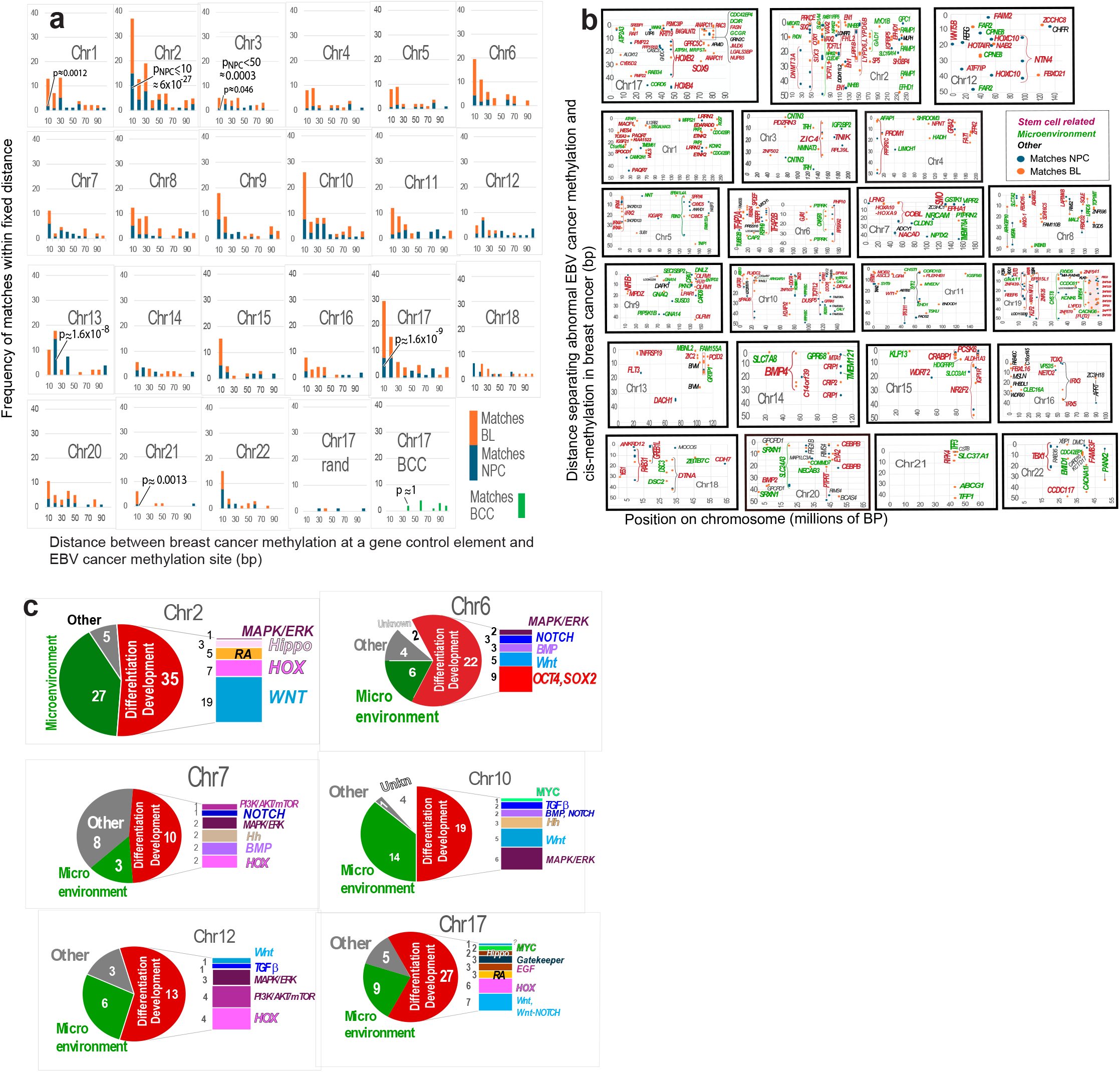
Matching methylation loci in EBV-cancers and breast cancers select for genes within stem-cell differentiation hierarchies. **a,** Proximity of abnormal methylation sites in breast cancer to those in EBV associated NPC and BL. Across most autosomes, breast cancer cis methylation of gene control elements sites clusters within 10 bp of NPC and BL sites. A chromosome 17 control shows almost no matches to a random distribution. The non EBV cancer BCC (green) does not exhibit the close positional concordance observed between breast cancer and the EBV associated cancers. **b,** abnormally methylated breast cancer regulatory regions align with abnormal methylation in known EBV cancers: NPC (blue) and/or BL (orange). Genes involved in differentiation (red), in regulating the microenvironment (green) or other functions (black) are also annotated. Many breast cancer regulatory sites coincide at identical or near identical positions in the EBV cancers, forming dense clusters. Chromosomes in the top row are described in Extended Data Note 1. On chromosome 19, the shaded region marks zinc finger gene matches, although data for this cluster are incomplete. **c,** The proportion of shared methylation events across functional categories. Bar charts indicate the differentiation pathways most frequently affected; gene counts appear to the left of each bar. On chromosome 6, shared methylation targets the pluripotency regulators OCT4 and SOX2. Most shared sites map to multiple pathways, but only predominant assignments are shown.

Skin basal cell carcinoma (BCC)^24^, a cancer unrelated to EBV, showed significant divergence, indicating that the observed similarities are not a generic feature of cancer methylomes (Fig. 1a).

These findings identify a conserved set of methylation events shared between breast cancer and EBV -driven tumors, suggesting a common upstream mechanism targeting gene regulatory elements.

### Shared methylation sites converge on lineage defining programs

To determine whether shared methylation represented preferential targeting of one or more specific upstream biological functions, loci were examined where breast cancer methylation^23^ overlapped NPC^14^, and BL^25^ loci. Shared sites preferentially mapped to promoters and enhancers of differentiation regulators (Fig. 1b). These loci govern plasticity, lineage commitment, and morphogen responsive patterning, and their disruption is known to promote stem-like cancer states^26–30^.

The patterns suggest that EBV-like methylation selectively expands stem-like states and supporting microenvironments across tumor types. Nearly half of overlapping loci involve progenitor cell identity, differentiation, or developoment genes (Fig. 1c, Extended Data Table 1).

### A conserved viral epigenetic strategy focuses on developmental controls

Overlapping methylated loci showed strong functional connectivity to genes annotated to developmental, stem-like, and lineage-defining programs. Among ∼350 of these genes, those shared across all three cancer types were predominantly related (40/58, 69%) to development, stem-like states, or differentiation pathways (Extended Data Table 1). The shared loci indicate that the same infection can target progenitors for both hematopoietic and epithelial cells. PANTHER GO over-representation analysis (Extended Data Table 2) found key developmental regulators were consistently hypermethylated across the methylated genes shared in the three types of cancers, suggesting broad interference with development / differentiation programs.

Matches (<10 bp) in abnormal methylation sites between breast cancer and either or both EBV-cancer chromosomes repeatedly focused on stem-cell differentiation hierarchies (Fig. 1b, c). Core regulators such as OCT4 and SOX3 were frequent targets, consistent with disruption of pluripotency networks^31,32^. In breast cancers, differential methylation of HOX regulatory elements, Wnt-associated genes, and other cell differentiation pathway genes was also shared, indicating substantial misprogramming of cell fate.

### Shared methylation loci dysregulate progenitor cell gene expression

Gene expression in breast cancers near methylation sites matching EBV cancers was significantly dysregulated across tumors (Fig. 2a–c). Strikingly, these genes fell into the established framework for epigenetic classes governing cancer stem cells (Extended Data Table 3), indicating that EBV-like methylation targets regulatory hubs with broad downstream consequences.

**Fig. 2.**
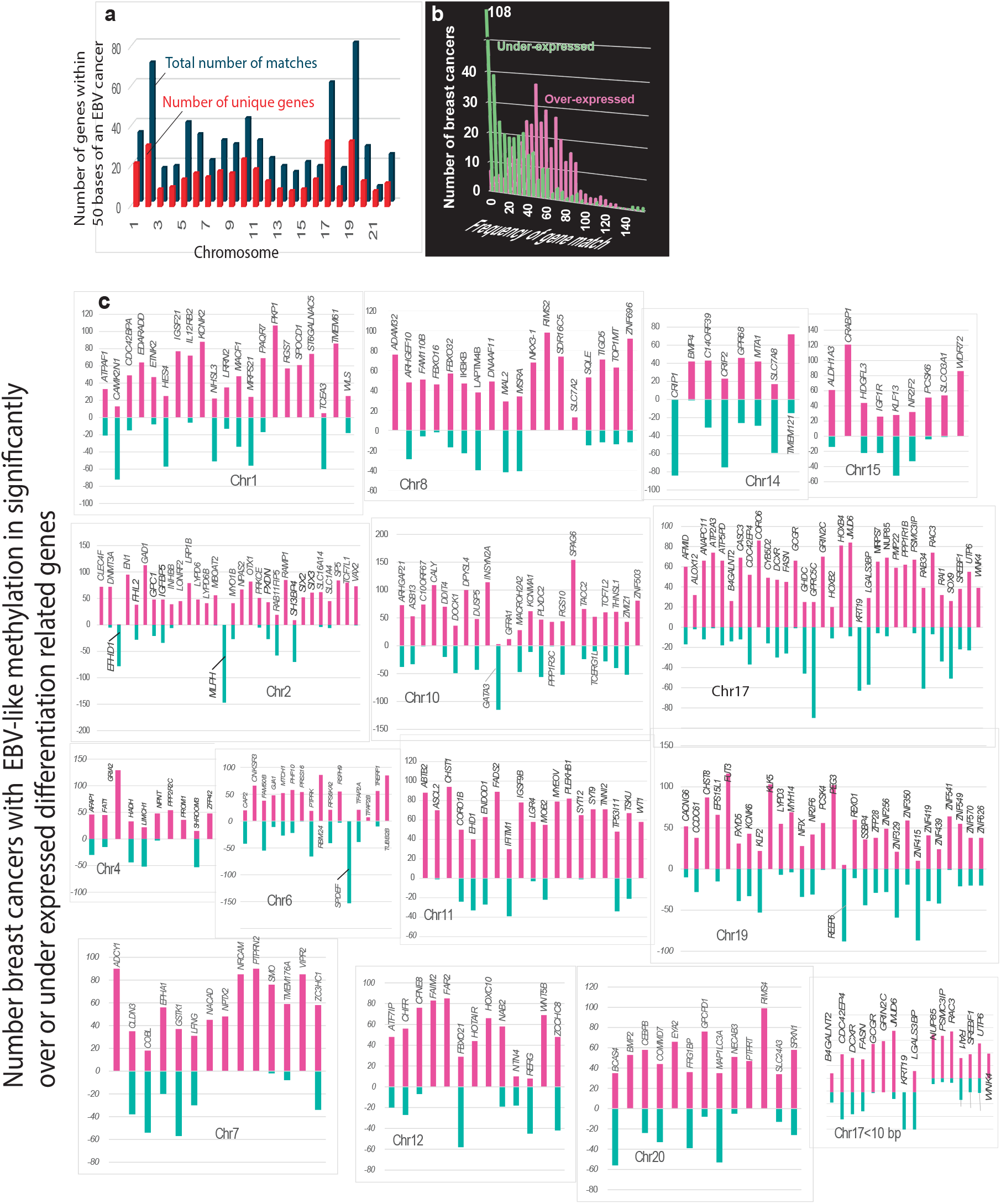
Robust changes in gene expression occur when abnormal methylation loci of breast cancer gene regulatory elements match those in EBV associated cancers. a, Abnormal methylation of regulatory regions of the same unique genes (red) can have multiple matches to abnormal methylations sites in EBV-cancers (blue). b, Genes controlled by EBV matching methylation sites are generally dysregulated in breast cancer. c, Breast cancer genes with abnormal methylation at NPC/BL matching sites (<50 bp) predominantly function in progenitor differentiation and show strong dysregulation (|z| > 2; red, over expression; green, under expression). The vertical axis indicates the number of tumors with abnormal methylation per gene. Many over expressed genes are not expressed in immune cells, making TIL based explanations unlikely. Genes are listed alphabetically within each chromosome panel. Numerous regulatory regions lie <10 bp from EBV methylated sites (example: chromosome 17). All genes near shared methylation loci with available expression data were significantly dysregulated.

Many affected genes were expressed almost exclusively in epithelial tumor cells, with negligible expression in infiltrating lymphocytes (Fig. 2). These tumor-intrinsic expression patterns are incompatible with infiltrating lymphocytes causing EBV-like methylation. Extended Data Table 4 reinforces this conclusion by comparing Human Protein Atlas expression data for twelve genes harboring methylation loci shared across all three cancers. These twelve genes perform biological functions irrelevant to lymphocytes, and their expression is absent or trivial in lymphoid tissues. Collectively, the data demonstrate that EBV-like methylation patterns originate within malignant epithelial cells, not from tumor-infiltrating lymphocytes, resolving a longstanding ambiguity in the relationship between EBV and breast cancer.

### EBV-like methylation converges across ER-positive and ER-negative breast cancers and is already established at the pre-Invasive stage

Luminal and basal breast lineages rely on distinct epigenetic programs, and tumors showed comparable EBV-like methylation overlap, suggesting a shared upstream mechanism (Fig. 3a). Both subtypes exhibited similar degrees of overlap with EBV-associated cancers. The two subtypes typically diverge sharply in their gene-expression programs so the shared methylation signature indicates a common upstream epigenetic mechanism.

**Fig. 3.**
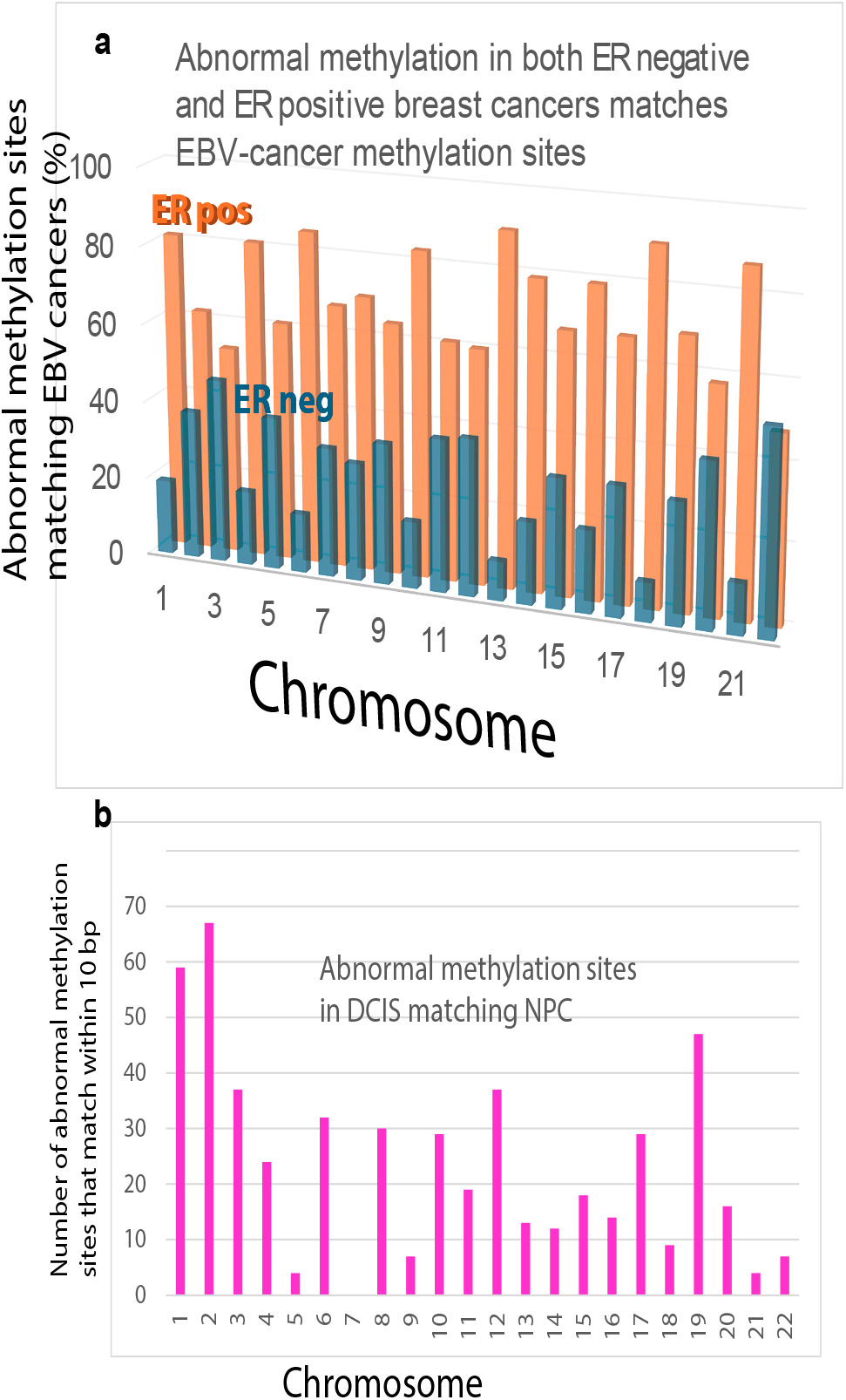
EBV-like methylation converges across ER-positive and ER-negative breast cancers and is already established in DCIS, the pre-invasive stage. a, Both ER-negative (typically basal type) and ER-positive (typically luminal type) breast cancers have similar amounts of overlap (10-52%) with abnormal cis-methylation sites that closely match cancers associated with EBV. Matches occur on all 22 autosomes and the shared sites are predominantly related to progenitor cell differentiation genes. b,.DCIS shows methylation changes that match NPC methylation positions within 10 bp for differential methylation sites differing from normal by >=0.1. The individual shared sites largely occur in sites related to differentiation or developmental programs.

To test whether shared methylation precedes invasive disease, methylation data from the early precursor lesion DCIS^34^ was examined. As shown in Fig. 3b, EBV-like deviations in methylation loci were already present in DCIS. The two adaptive immune terms (Extended Data Table 5) reflect a strongly enriched change focused on the adaptive immune response but driven by only about 4-5% of the genes. Developmental pathways form the dominant epigenetic signature involving 79-110 genes. Most epigenetic alterations involve broad, coordinated disruption of tissue identity and morphogenesis programs. DCIS thus shows large scale epigenetic reprogramming of genes that maintain epithelial architecture and differentiation. Because it precedes development of invasive cancer, EBV-like methylation therefore is unlikely to be caused by increased susceptibility to EBV due to the breast cancer.

To determine whether these epigenetic signatures reflect a lasting imprint of prior EBV infection and an intrinsic feature of infection that is not exclusive to tumors, data was examined from immortalized normal oral keratinocytes (NOKs) that had been infected with EBV and subsequently cured. These non-malignant cells retained DNA methylation patterns strongly correlated with those observed in breast cancer (r = 0.93), clustered near stem-cell regulatory genes across 14 chromosomes (Fig. 4). Additional genes methylated in the cured NOKs that were not methylated in breast cancer were primarily related to the microenvironment (Extended Data Table 6).

**Fig. 4.**
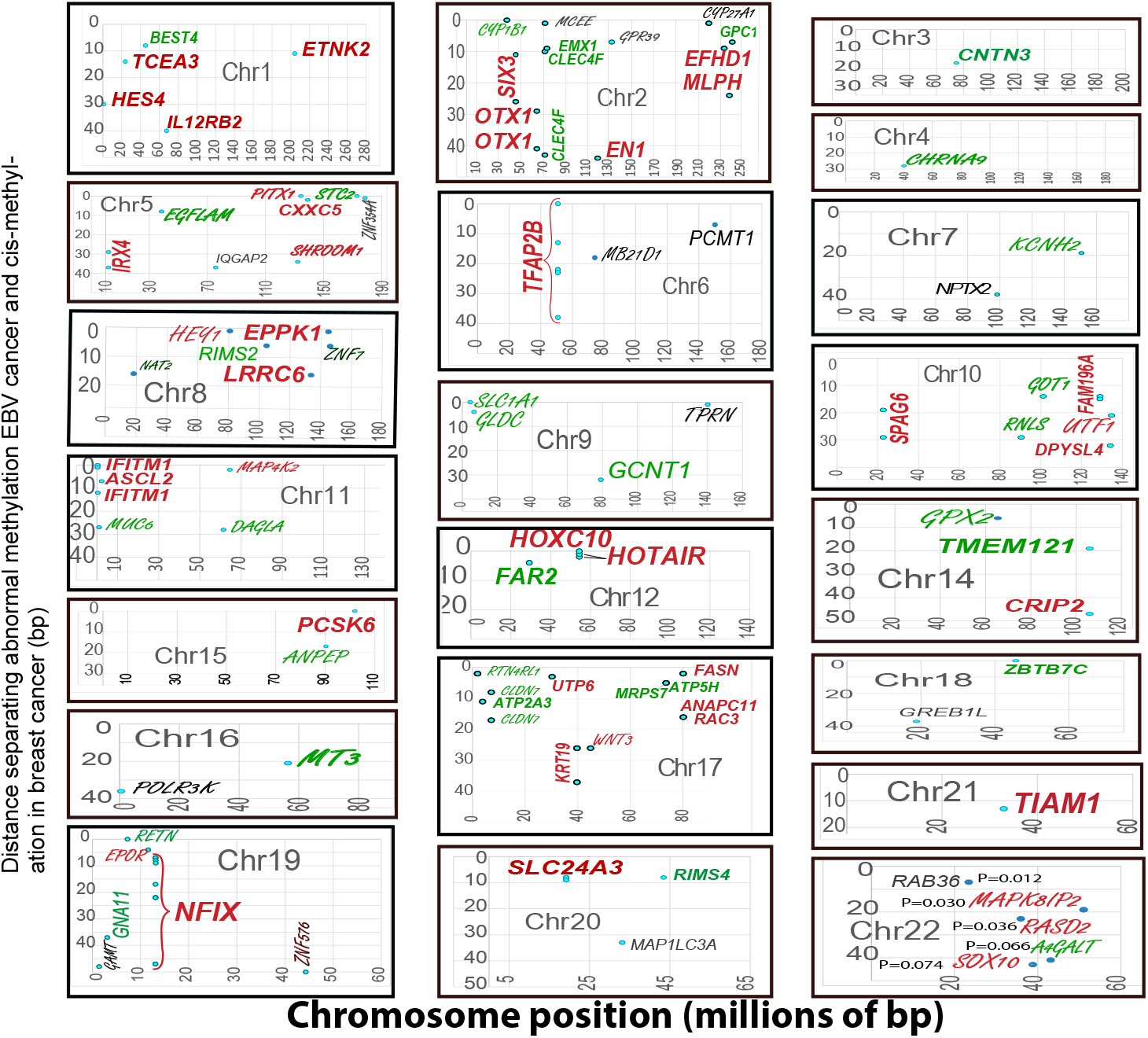
EBV imprints stable methylation changes in non-malignant NOK cells near elements that affect cell differentiation and resemble epigenetic changes in breast tumors. Methylation positions in NOK cells with a cured prior EBV infection that matched either NPC or BL were compared with abnormal cis-methylation in breast cancers. Most shared sites involve gene controls for progenitor cell–like genes (red) or microenvironment-related genes (green); other matched genes are shown in black. Genes methylated in EBV-exposed keratinocytes but not in breast cancers appear in script font. Chromosome 22, a relatively short chromosome was compared to chance occurrence, which showed most matches are unlikely due to chance.

To confirm that this pattern is EBV-specific rather than a general feature of epithelial cell transformation, the cured keratinocyte methylation profile was compared to that of basal cell carcinoma (BCC)^24^, a skin cancer with no known EBV association. The negligible overlap between methylation sites in cured NOKs and BCC (Extended Data Fig.1), demonstrates that the shared signature is not simply a product of epithelial origin or carcinogenic stress, but reflects a distinct EBV-driven epigenetic program.

Collectively, these results indicate that EBV imprints stable methylation changes on epithelial cells that persist after viral clearance, and cluster near progenitor cell regulatory genes. The persistence of some of these changes (Fig. 4) supports an origin intrinsic to infection. Some changes mirror the epigenetic landscape observed in both ER-positive and ER-negative breast tumors. These findings are consistent with EBV-linked epigenetic reprogramming beginning at least as soon as DCIS, and acting on luminal progenitors across both lineages. This chronic, lasting disease misprogramming results from sustained tissue-specific pressures (including inflamation), which precede and may actively drive subsequent malignant progression.

### Corroborating evidence for an antiviral response

To test whether breast cancer genomes showed evidence of an antiviral response, indicators of antiviral activity were assessed: Interferon-stimulated genes (ISGs), retroelements, and piRNA–PIWI pathway activation. The results showed that ISGs formed a highly coordinated antiviral module, while endogenous retroelements showed no correlation with ISG expression (Extended Data Figs. 2a, b). This distinction suggests the immune response is being driven by something other than the cell’s own genetic elements. Notably, HLA class D antigen-presentation genes (which help the immune system recognize and respond to infected cells) tracked closely with ISG expression (Extended Data Fig. 2b).

PIWI proteins, PIWIL2 and PIWIL4 were highly over-expressed compared to PIWIL1 and PIWIL3. This pattern is consistent with activation of the antiviral piRNA epigenetic machinery, a defense system that cells use to silence viral and repetitive genetic elements (Extended Data Fig. 2c). Extended Data Fig. 2d shows that in breast cancers antiviral innate and adaptive immune response pathway expression rises and falls in coordination.

### Alternative Explanations

Multiple alternative explanations for the breast-cancer methylation and immune activation patterns were evaluated and found incompatible with the data. In brief, the affected loci are not expressed in infiltrating B-cells; the coordinated rise and fall of ISGs argues against a non-viral origin; endogenous retroviral expression does not track with ISG signatures; age-related or replication-clock drift^35^ showed almost no overlap with the relevant control-element sites (Extended Data Fig 3); general cancer-associated methylation fails to match patterns from non-EBV skin basal cell carcinoma; and statistical testing rejects chance overlap with high confidence. Together, these analyses rule out these conventional explanations and reinforce the idea that shared methylation anomalies represent a distinct underlying process.

### EBV-linked methylation patterns extend to the neurologic disease MS

To determine whether EBV-associated epigenetic misprogramming extends into tissues with chronic diseases that are not malignant, data from multiple sclerosis (MS) patients was examined. EBV involvement in MS is well established ^17^, yet MS arises in a fundamentally different non-epithelial cell context within a restricted-access CNS compartment.

The first comparison (Fig. 5a) revealed close matches to NPC for abnormal methylation sites in pathology-free MS brain tissue^36^ . Although MS is driven by an EBV peptide that provokes autoimmune demyelination^18^, the non-pathologic MS brain tissue being compared had no visible overt inflammation or demyelination on staining. Results identified HOXD3 and NDRG4 as shared abnormal methylation sites with relationships to cell differentiation and identity (Extended Data Table 7). HOXD3 is an especially high-value EBV target for manipulating cell differentiation because it is a plasticity controller in multiple tissues^37,38^.

**Fig. 5.**
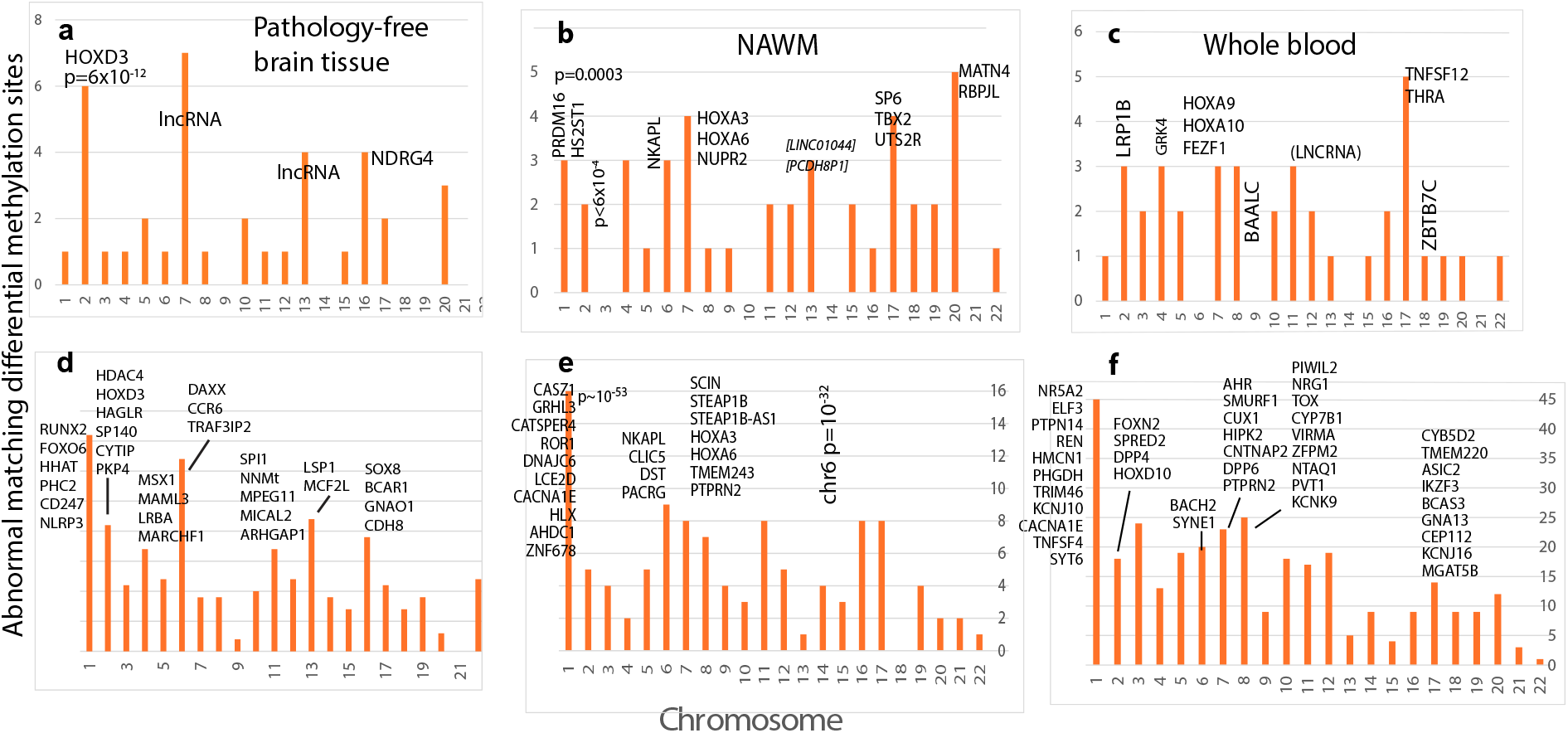
Tissues affected by epigenetic misprogramming include the CNS. Abnormal methylation in MS samples associated with NPC around the likely site of primary EBV infection shows positional matches to differential methylation sites. The top row graphs the numbers of NPC methylation sites on each autosome that coincide with differential methylation in MS samples: a, pathology free brain tissue b, NAWM, and c, blood. Many genes at these shared loci contribute to stem cell differentiation pathways (Extended Data Tables 7–9), and representative individual genes are shown on their chromosomes. Methylation data from 60 breast cancers was next compared to the same MS microarray data (bottom row): d, pathology free tissue, e, NAWM, and f, blood. Differentiation related functions of the indicated genes are summarized in Extended Data Tables 10–12, Many of the matches shown in panels a-f were exact positional matches. Representative p values from panels a-f indicate that observed agreement is unlikely to occur by chance.

To confirm the relationships to cell differentiation, data for glial cells within NAWM from MS patients was next compared to the same NPC data. Like the pathology-free cohort, the NAWM samples did not have detectable CNS inflammation^39^, and they still showed EBV-like methylation at differentiation-related loci (Fig. 5b, Extended Data Table 8). In NAWM, aberrant methylation in HOXD3 and HOXA3 overlapped NPC loci and provided clear examples of targeted selection for interference with differentiation. The two genes are paralogs that together pattern the pharyngeal region, including the thymus.

The oropharyngeal / nasopharyngeal lymphoid tissue of Waldeyer’s ring is the primary site of EBV infection and its lifelong latency reservoir. This tissue sits directly within the pharyngeal field patterned by HOXA3/HOXD3 during development. HOXA3 initiates thymus organ identity^40^; while HOXD3 acts with HOXA3 to support development of thymic epithelial cells, which train developing T-cells to distinguish self from non-self. Even though the specific paralog marked differs by cohort, the HOXA3 signature seen in the NAWM cohort implicates the same thymic developmental pathway as HOXD3 in pathology-free brains.

Some methylation abnormalities in MS spill over into blood^41^ and these were assessed next. Genes related to cellular differentiation pathways are strongly represented (Fig 5c, Extended Data Table 9). Methylation sites in the blood cohort matching NPC include HOXA9 and HOXA10. HOXA9 and HOXA10 are consistently upregulated in NPC transcriptomic datasets and both contribute to the same posterior developmental module that promotes matrix remodeling and migratory behavior relevant to NPC progression^42,43^.

To test whether these results applied to a completely different tissue, breast cancer methylation patterns different from adjacent normal tissue^44^ were next compared to the three independent MS cohorts. These comparisons again highlighted abnormally methylated differentiation related genes (Fig 5d-f, Extended Data Table 10-12). To illustrate the potential effects from the many overlaps found, HOXD10 on chromosome 2 is frequently dysregulated in breast cancer and contributes to invasive progression through cytoskeletal remodeling and motility pathways (Fig. 5f, Extended Data Table 12). Control comparisons to data from CD4 positive T-cells^45^ of MS patients showed only rare agreement across all autosomes. (Extended Data Fig. 4).

The same or closely related HOX patterning genes turn up epigenetically altered in NPC, a cancer arising at the probable initial site of infection, in MS whose blood brain-barrier-disrupting-lymphocytes plausibly originate in that same lymphoid niche, and in cancer of the breast. In the public data used, postmortem MS patient samples had been sorted to exclude peripheral immune cells, including lymphocytes^39^. Thus, EBV-positive non-inflamed tissues likely represent an early, smoldering stage of CNS viral involvement. Together, these cross-tissue methylation patterns indicate that EBV-associated epigenetic misprogramming extends beyond malignant settings into non-inflamed CNS tissue and spills into peripheral blood. Prior direct tissue-level evidence supports this broader model: EBV-infected breast epithelial cells form mammary tumors when transplanted into immunodeficient mice^20^, and EBV-related antigens are detectable within glial cells in active MS lesions^46^. These primary observations provide additional support for the idea that EBV selects for progenitor-level changes across multiple diseases, providing a basis for the mechanistic interpretation in the Discussion.

## Discussion

The results suggest EBV changes markings within stem-cell hierarchies in multiple tissues. These changes are then propagated and selected for as descendant cells differentiate along divergent paths. Thus while different studies and methodologies may highlight different sets of genes, they are capturing the same underlying biological process connecting several previously unrelated diseases. Methylation abnormalities that align at virtually identical CpG coordinates across NPC, BL, breast cancer, and MS (p-values near 0) are difficult to reconcile with models in which each disease arises through independent, tissue-specific mechanisms. Instead, these findings point to a conserved upstream event: epigenetic reprogramming of progenitor chromatin that precedes and links context-dependent pathology. This implies that EBV infects long-lived, self-renewing progenitors in whatever lineage it enters, often as slightly different variants in different compartments of the same host^47^. EBV latency proteins (e.g., LMP1/EBNA3C) recruit DNMT/TET machinery to reprogram infected lineage regulatory loci. NPC and Burkitt lymphoma may therefore result from the same disease strategy but in different contexts. The tactic of hijacking host epigenetic machinery is the same even as the target cell, entry receptor, and specific CpG sites remain lineage-specific. Cancer in the breast has been reported as a metastatic site from NPC^22^, but it was never clear that a shared molecular program linked the two diseases.

Perhaps the most striking aspect of this convergence is that it spans tissues with fundamentally different developmental origins, cellular composition, and clinical presentation: nasopharyngeal epithelium, B-lymphocytes, breast epithelium, and CNS glia.

Affected loci are consistently enriched at progenitor-biased regulatory elements governing lineage commitment (including HOX clusters, WNT/BMP components, and stem-cell factors such as OCT4, SOX2, and SOX9) rather than at mature effector genes. This enrichment again suggests that the chromatin architecture enforcing differentiation commitment, rather than any single tissue-specific program, is selected for by the EBV epigenetic strategy.

The breast cancer findings help resolve a long-standing controversy: the EBV-like signature is expressed in malignant breast epithelial cells themselves, and does not require infiltrating lymphocytes. It is already detectable in DCIS, indicating a tumor-intrinsic origin that precedes invasive disease so invasive disease does not cause EBV-like signatures. That cured, non-malignant keratinocytes retain remnants of this signature with high fidelity (r = 0.93) shows that the epigenetic scar is self-perpetuating. The virus need not persist once its epigenetic instructions are written. A parallel pattern in non-inflamed MS brain tissue, where glial methylation overlaps EBV-associated cancer loci, extends this principle to the CNS and implicates progenitor level disruption rather than lymphocyte mediated injury alone as an early step in disease.

Together, these results reframe the temporal relationship between EBV infection and chronic disease: the critical pathogenic event may be early epigenetic misprogramming within stem-cell hierarchy rather than viral persistence, reactivation, or inflammation. The consequences propagate silently through cell generations until secondary insults convert latent vulnerability into overt disease. Given that EBV infects over 90% of adults worldwide, often during life stages of active progenitor proliferation, this mechanism may account for a larger share of chronic disease than currently appreciated. Stem-cell vulnerability within differentiation pathways emerges as a mechanistic common denominator and a biomarker across diverse, apparently unrelated diseases, pointing to early epigenetic correction as a potential avenue for intervention long before clinical disease appears.

## Methods

### Breast cancer patients and methylation data

Clinical and molecular data for 1,904 female patients with primary breast cancer were obtained from the METABRIC cohort through the original publications 48 and cBioPortal. The cohort included 1,459 estrogen receptor (ER)-positive and 445 ER-negative tumors. Tumor grade distribution was 165 grade 1, 740 grade 2, and 927 grade 3 cases.

Most tumors were ductal carcinomas (n = 1,454). Patient ages ranged from 22 to 96 years, with the majority (1,200/1,904) between 47 and 72 years. Available annotations included clinical characteristics, tissue procurement metadata, genome-wide expression profiles, copy-number aberrations, and somatic point mutations^48,49^.

DNA methylation data were from published whole genome reduced representation bisulfite sequencing of 1,538 breast tumors and 244 adjacent normal tissues within the METABRIC cohort 23 . Promoter regions were screened for cis Pearson correlations methylation sites and gene expression on the same chromosome; trans effects were excluded.

Cis-acting sites were defined as those showing the strongest negative correlation with their corresponding genes across >50 tumors (FDR<0.05). Additional microarray-based methylation data from 60 matched tumor–adjacent normal pairs in a separate multi-institution study 44 were incorporated. Finally, methylation and expression data for 1111 primary tumors in 1095 different patients with 7 metastatic cancers were downloaded from TCGA.

### NPC patients and methylation data

NPC methylation data were derived from whole genome bisulfite sequencing of biopsies from 15 sporadic NPC patients and 9 matched non-tumor adjacent tissues 14. All tumors were primary NPCs (stage II, n=3; stage III, n=7; stage IV, n=5). Patients ranged from 38–82 years (13 males, 2 females), and 11 were survivors. Differential methylation was defined as the average difference between NPC and matched normal tissue. In total, 16,910 differentially methylated regions (DMRs) exhibited absolute methylation deviation >0.2, and 6036 were <500 bp long with average differential methylation of -0.336 to 0.463. Three NPC samples were globally hypomethylated and 12 globally hypermethylated. Results were validated in an independent cohort of 48 NPC patients.

### BL patients and methylation data

BL methylation data came from a published cohort of 13 patients (ages 3–18; 11 males, 2 female) 25. All tumors harbored IG–MYC rearrangements and exhibited differential hypermethylation correlated with gene expression. More than two-thirds of the DMRs were <500 bp, with lengths varying from 8 to 5,160 bp. Reference DNA was isolated from non-neoplastic germinal center B-cells in tonsils of four donors (ages 13–30). The present analysis used 42,381 hypermethylated DMRs across the 22 autosomes. Average methylation levels in germinal center B-cell DNA were subtracted from corresponding BL values, retaining only regions showing ≥20% differential methylation.

### Transiently infected non-malignant NOK methylation data

Source data 50 were generated from hTERT immortalized NOKs infected by 24 h coculture with anti-IgG–induced Akata BL cells. After B cell removal, infected NOKs underwent 10 passages under antibiotic selection followed by 10 passages without selection to generate transiently infected populations. Single-cell clones were isolated by flow cytometric sorting; EBV status was confirmed by EBER in situ hybridization.

Uninfected parental NOKs and plasmid vector controls were cultured in parallel. All clones were authenticated by DNA fingerprinting. Methylation profiling was performed using reduced-representation bisulfite sequencing.

#### BCC methylation data

BCC data came from 16 biopsy specimens (11 women and 5 men, ages 55-89) comprising 8 nodular and 8 sclerodermiform BCCs 24. Tumor sites included face, shoulder, head, ear, lip, nose, and pectoral areas. Whole genome methylation profiling used Infinium Methylation EPIC Bead Chips.

### MS patients and methylation data

MS samples were from post-mortem (<31h) frontal lobe brains lacking inflammatory infiltrates or plaques^36^. Available Illumina methylation data comparing methylation levels in MS samples against matched controls were analyzed. Statistical significance was assessed using model-based likelihood-ratio test (LRT) p-values combined using Fisher’s method to generate an aggregated significance statistic for each CpG. Multiple-testing correction used the Benjamini–Hochberg false discovery rate (FDR). CpGs were considered significantly differentially methylated if they met both criteria: Fisher’s method p-value < 0.05 and FDR q-value < 0.05. NAWM data were obtained from 8 MS patients and 14 non-neurological disease controls^39^. Blood data was from 70 MS patients (53 female and 17 male)^41^.

### Data harmonization

Potential sources of error included bisulfite non-conversion, coverage differences, and methodological bias across platforms (WGBS, RRBS, and Infinium 450K/EPIC arrays), as well as cell-type composition and batch effects. These technologies do not produce fully compatible or interchangeable datasets, and cross-platform comparisons were therefore restricted to features that can be reliably aligned.

To harmonize datasets, all genomic coordinates were first mapped to a common genome build (GRCh37/hg19 or GRCh38) using UCSC LiftOver, retaining only uniquely mapping loci. Methylation values were reduced to average beta differences (β_tumor – β_normal), and non-significant sites were removed using platform-specific thresholds. Differentially methylated regions (DMRs) were called independently within each platform and only overlaps in genomic coordinates or affected genes were compared across datasets. Gene-expression comparisons used Z-score normalization.

### Calculations of matching abnormally methylated positions

Distances between abnormally methylated positions in breast cancer and EBV associated cancers (NPC or BL) were computed as the minimum absolute difference between the start, end, and midpoint coordinates of each breast cancer cis hypermethylated segment and EBV associated DMR coordinates. A >20% deviation from normal cell methylation was applied across the full length of each segment. An example of the Excel spreadsheet formula (Fig. 1) for minimum distance calculation was: =MIN(ABS(D2-$K$2:$M$1532),ABS(E2-$K$2:$M$1532),ABS(F2-$K$2:$M$1532)), where columns D through F contain breast cancer cis-hypermethylation positions and columns K through M contain differential hypermethylation positions in EBV cancer (either NPC or BL).

Statistical significance was assessed using Poisson probabilities, with λ defined as the expected number of random overlaps. Poisson distributions were evaluated in Python 3.13. Hypergeometric tests were also applied, for example to assess overlap between MS associated and breast cancer associated CpG sites on chromosome 1. Using Illumina EPIC probe backgrounds (MS: 700,482 probes; BC: >850,000 probes), the expected overlap was ∼1.23 sites. The observed 16 positional matches were highly unlikely to occur by chance (Poisson approximation p ≈ 4.1 × 10^-^¹³). Even if spatially clustered probes at 26.2 and 227.6 MB were collapsed into 12 independent loci, enrichment remained significant (p ≈ 8.0 × 10^-^⁹).

### Calculation of frequency of close agreement

Frequencies of agreement between breast cancer and EBV-cancer were calculated in Excel after normalizing NPC data to the same total numbers of cancers as BL data (15 NPC vs. 13 BL).

#### Gene-feature mapping

All genomic coordinates were taken directly from the original publications and used directly. For MS related gene comparisons, genomic coordinates were mapped to gene features using GENCODE v50 (GRCh38). GTF annotations were imported into R using the rtracklayer package to obtain gene level metadata including gene_id and gene_name. Genomic ranges corresponding to the input coordinates were constructed with GenomicRanges, and overlaps with annotated gene bodies were identified using findOverlaps. Gene identifiers from the TxDb object were matched to GENCODE gene metadata to retrieve gene symbols. Coordinates overlapping multiple genes or antisense transcripts were retained and reported. Analyses were performed in R version 4.x.

### Gene ontology analysis

Pathway enrichment analysis of breast cancer differentially methylated positions^51^ (DMPs) was performed using the PANTHER over-representation test

### Breast cancer gene expression data

Gene expression data came from the METABRIC study 48 using the Illumina HT12 v3 microarray. Genes with expression z scores ≥2 SD from the mean (p < 0.05) were identified and compared with stem-cell and differentiation related genes located within 50 bp of differential methylation sites in EBV-related cancers.

### Comparisons of distributions

Normality was assessed using StatsDirect. Hypermethylation distributions consistently deviated from normality and were evaluated by Mann–Whitney nonparametric tests. Correlations between breast cancer and EBV-associated epigenetic modifications were assessed by Kolmogorov–Smirnov and Spearman’s rank tests. Frequency plots were generated to visualize coincident regions.

#### Resource Availability

Datasets used are well-documented and freely available from original sources or the author on reasonable request.

### Use of generative AI tools

Generative AI tools were used to assist with coding and text editing. All AI generated code and edited text was manually inspected, corrected when necessary, and validated by the author, who takes full responsibility for the final analyses and manuscript content.

## Data Availability

All data produced in the present study are available upon reasonable request to the author

## Extended Data

**Extended Data Fig. 1.**
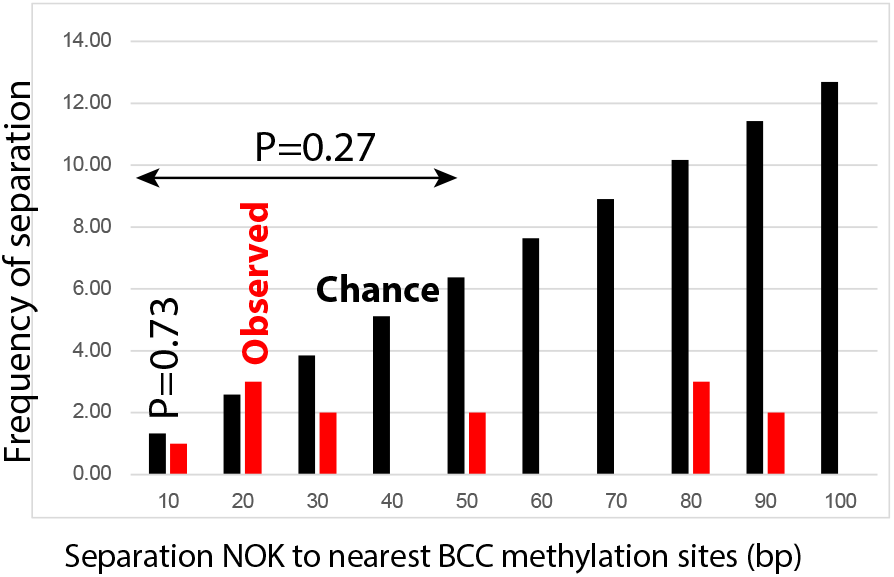
Differential methylation is unlikely due to chance. The frequency of differentially methylated genes in normal oral keratinocytes that have recovered from an EBV infection (NOK)s within 50 bp of methylation sites in basal cell carcinoma of the skin is generally less than expected by chance. Observed frequency data is in orange and frequencies expected by pure chance are in black.

**Extended Data Fig. 2a.**
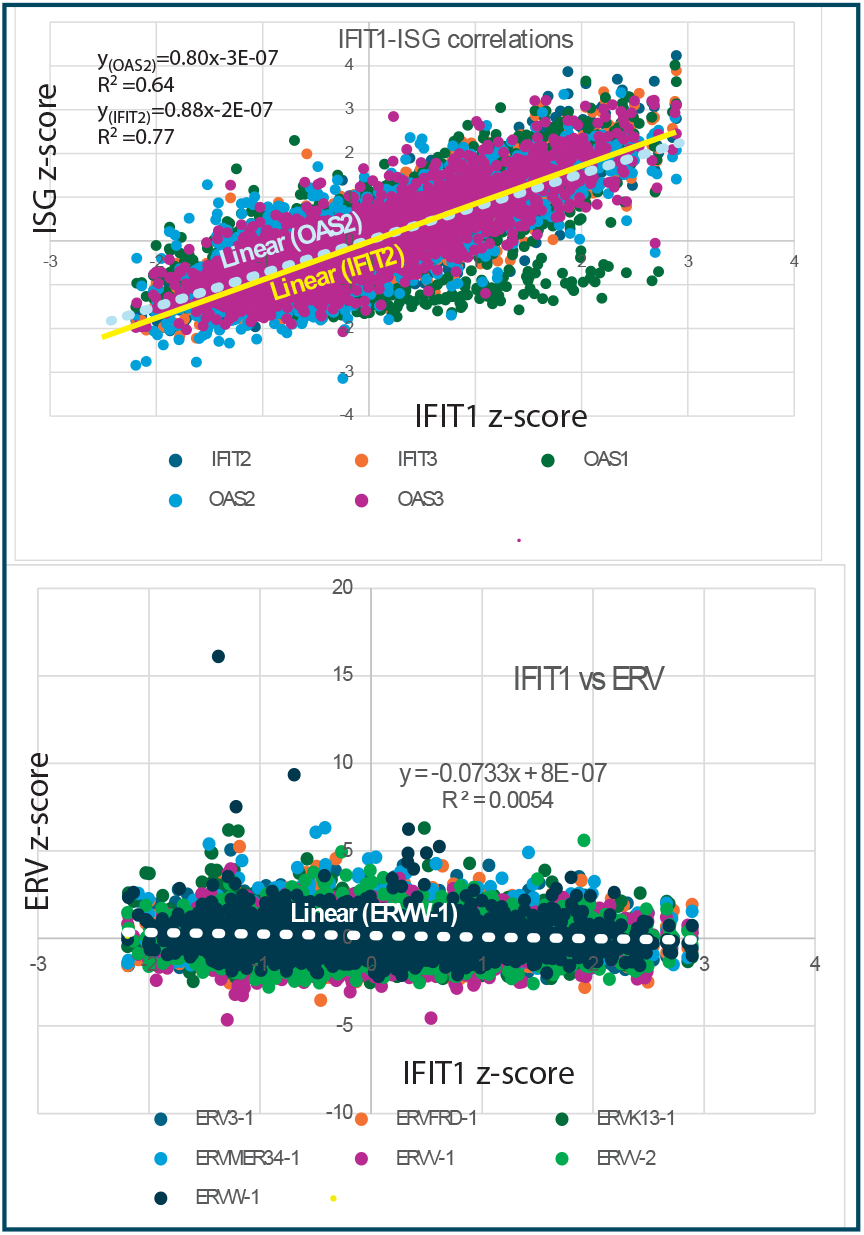
Evidence for an antiviral response in breast cancer. IFIT1 correlates strongly with other ISGs but has no coherent relationship to ERVs expressed in breast cancers. ERW-1 is a functional Env gene encoding human protein syncytin-1 ERW-2 is a non-expressed HERV-W fragment.

**Extended Data Fig. 2b.**
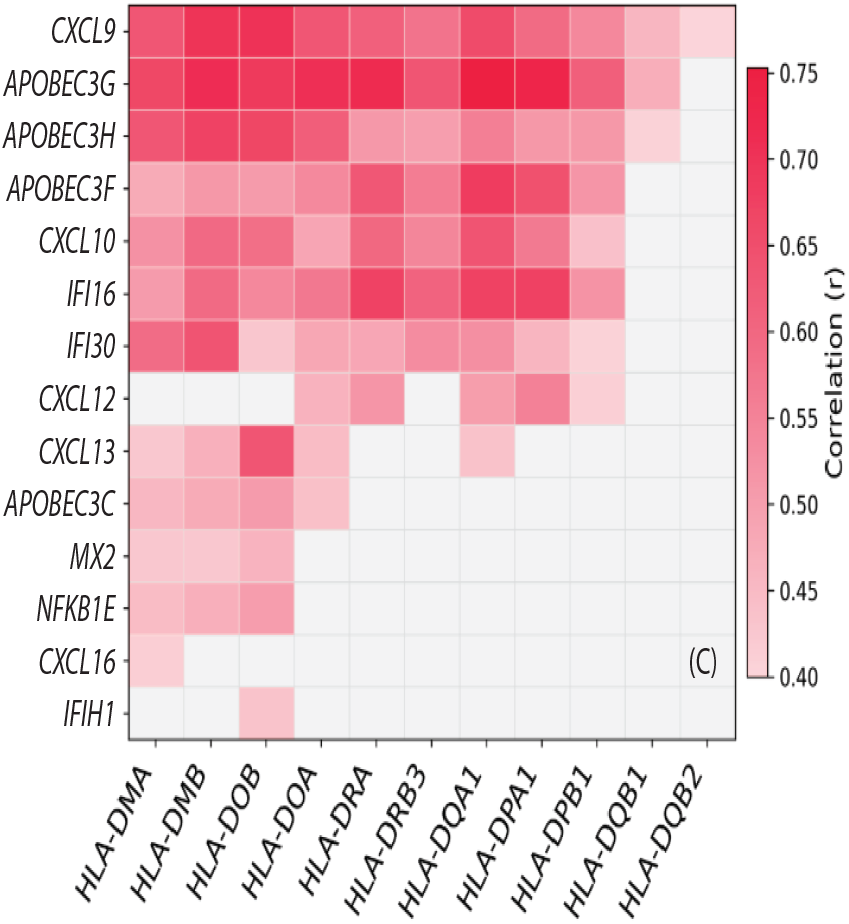
Breast cancers have been responding to an exogenous infection. Interferon stimulated genes strongly correlate with HLA-D antigen presentation genes.

**Extended Data Fig. 2C.**
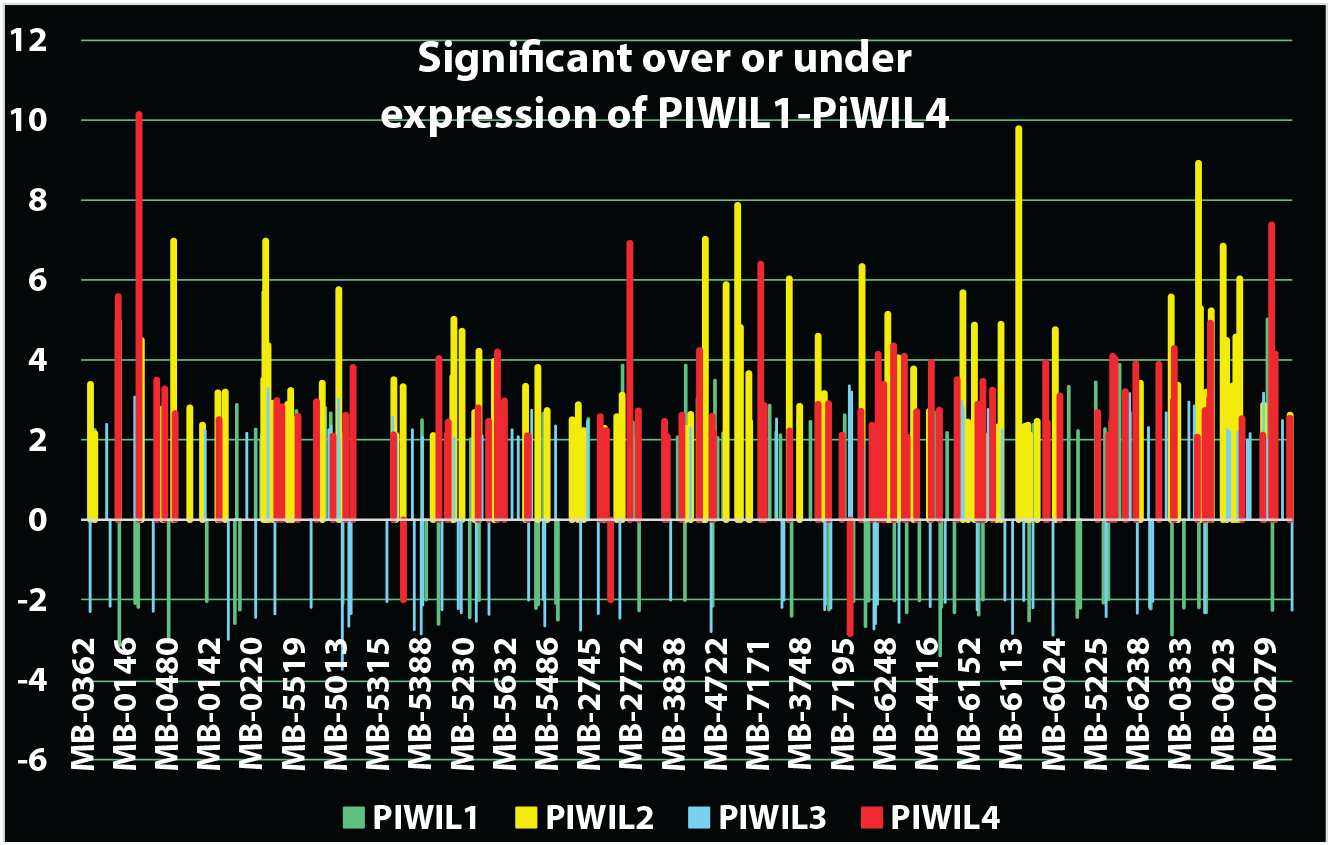
Dysregulation of the piRNA system in breast cancers showing abnormalities in an antiviral response system. PIWIL1-PIWIL4 genes with z>=|2| are shown. Overexpression occurs predominantly for PIWIL2 and PIWIL4. PIWIL1 and PIWIL3—linked to differentiation-associated piRNA pathways—are frequently suppressed, consistent with tumors avoiding developmental programs that oppose stemness or increase viral-mimic stress.

**Extended Data Fig. 2d.**
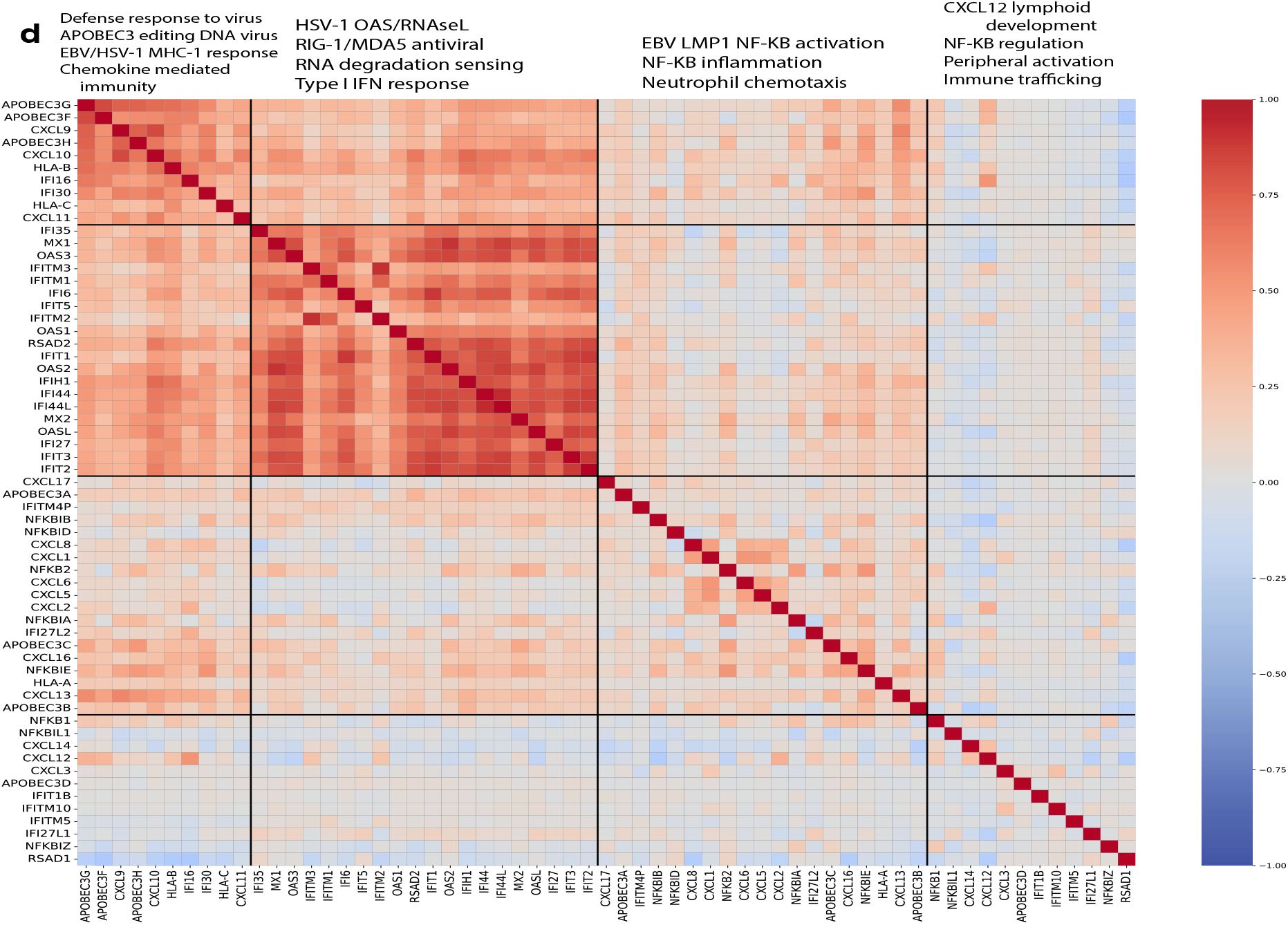
Further evidence for an antiviral response in breast cancers. Antiviral innate and adaptive immune pathways expressed in breast cancer typically engaged during viral infection have correlated expression. Abnormally expressed antiviral modules 1 and 2 are co-expressed together. Alternate explanations not involving a virus have no support based on available medical information.

**Extended Data Fig. 3.**
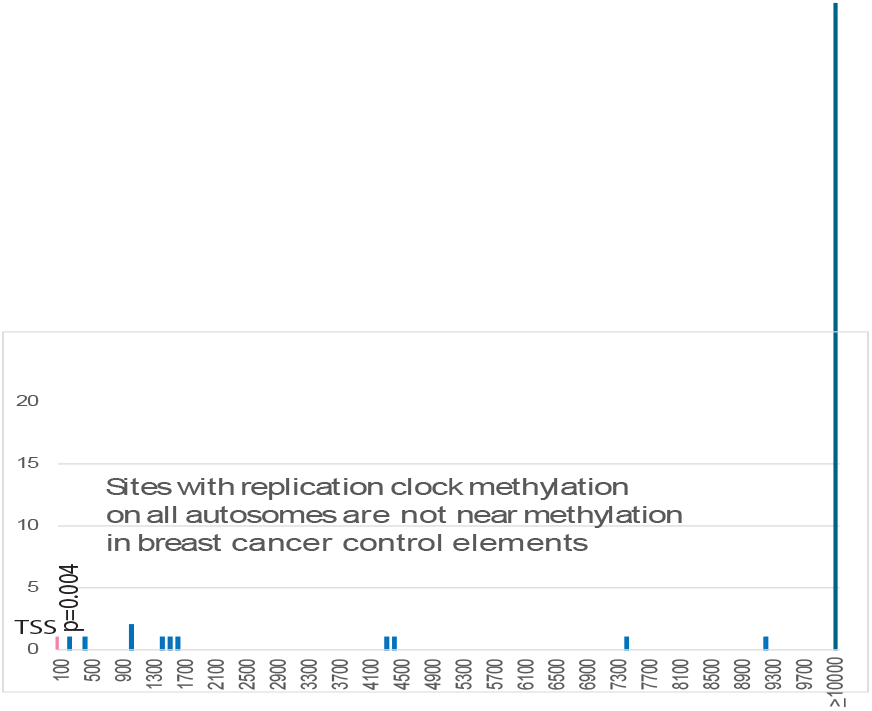
Most differential methylation in breast cancer is not caused by epigenetic drift in aging. Differential methylation of breast cancer control elements only rarely matches replication clock sites in old vs young cells. The nearest distances between methylation in breast cancer control elements and methylation more characteristic of old cells were relatively large on all chromosomes. This suggests agreement in these patterns are not primarily attributable to passive epigenetic aging. On chromosome 11 a map position that was more heavily methylated in old cells matched within 174 bp(p=0.0074). When compared to transcription start sites the same match was reduced to 12 bp. However most replication clock sites were >10000 bp removed from a breast cancer methylation site.

**Extended Data Fig. 4.**
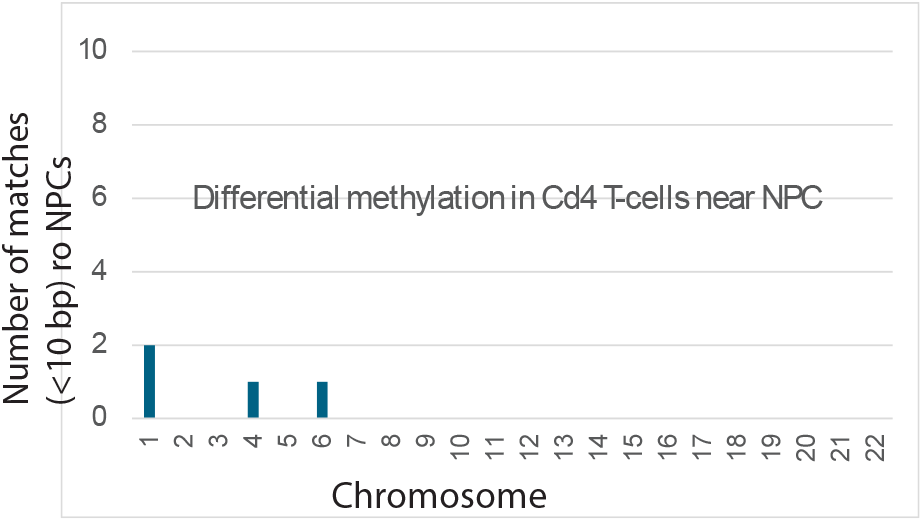
Differential methylation that matches NPC does not affect helper T-cells in MS (negative control). The frequency of differential methylation sites in CD4 positive T-cells from MS brains that match differential methylation sites in NPC. The CD4 T-cells came from the 2017 Maltby study of 28 MS patients with relapsing-remitting MS.

## Extended Data Note 1

In Fig. 2, chromosome 17 was examined first because it encodes BRCA1 and ERBB2 (HER2) genes, both significant in breast cancer. In the breast cancer cohort, chromosome 17 had 405 gene entries with abnormally methylated cis-regulatory sites with 85% <=200 bp long. As shown in Fig. 2, comparisons of abnormal methylation sites on chromosome 17 in breast cancer [1], NPC [2], and BL [3] converged on promoters or enhancer regions of genes associated with stem-like or differentiation / developmental lineage programs.

For instance, differential methylation sites in the three different cancers were found at regulatory regions of SOX9. SOX9 counterbalances the pluripotency factor SOX2. Together SOX9 and SOX2 shape cancer cell plasticity and metastasis [4]. SOX9 also participates in the NOTCH signaling network governing mammary cell self-renewal and lineage choice [5]. KRT19, a NOTCH signaling control point [6] , was methylated within three bases of the same regulatory positions in breast cancer and BL. Breast cancer and EBV-associated tumors shared hypermethylated clusters within regulators of the HOXB locus, including regions controlling HOXB2 and HOXB4. These blocks overlapped regulatory elements characteristic of progenitor cell chromatin states [7]. HOXB genes have documented roles in both breast and EBV-associated cancers [8, 9].

Finding SOX9, KRT19, and HOX genes with methylation at nearly identical regulatory positions in breast and EBV-driven cancers suggested that EBV-induced methylation had occurred in breast stem-like or progenitor cell differentiation programs, potentially disrupting them. Fig. 1 hints at this possibility: children with BL consistently exhibited more abnormal EBV-matching methylation than adults with NPC, consistent with the well documented agerelated decline in stemcell numbers and lineage potential. This pattern could merely reflect differences in tumor cell biology, but it motivated broader analyses on additional breastcancer autosomes.

On chromosome 17 (Fig. 2, 3), ∼66% of abnormally methylated cis-regulatory positions in breast cancer coincide with EBV cancer methylation loci. On chromosome 2, genes with altered controls in both breast and EBV-cancers, (including HOX, SIX2, SIX3, VAX2, and EN1), have strong developmental and stem-like cell connections (Fig. 2, 3). HOX genes set broad positional identity [10], SIX, VAX, and EN genes refine regional patterning and morphogen gradient responses essential for tissue homeostasis [11] . Chromosome 12 results include an RNA-based mechanism. The long noncoding RNA HOTAIR, a 2.2 kb antisense transcript, represses HOXD cluster transcription by recruiting chromatin modifying complexes [12]. Across all autosomes, 47% of coinciding genes participate in differentiation / developmental pathways, with some related to core stem cell genes.

---

**Extended Data Table 1.**
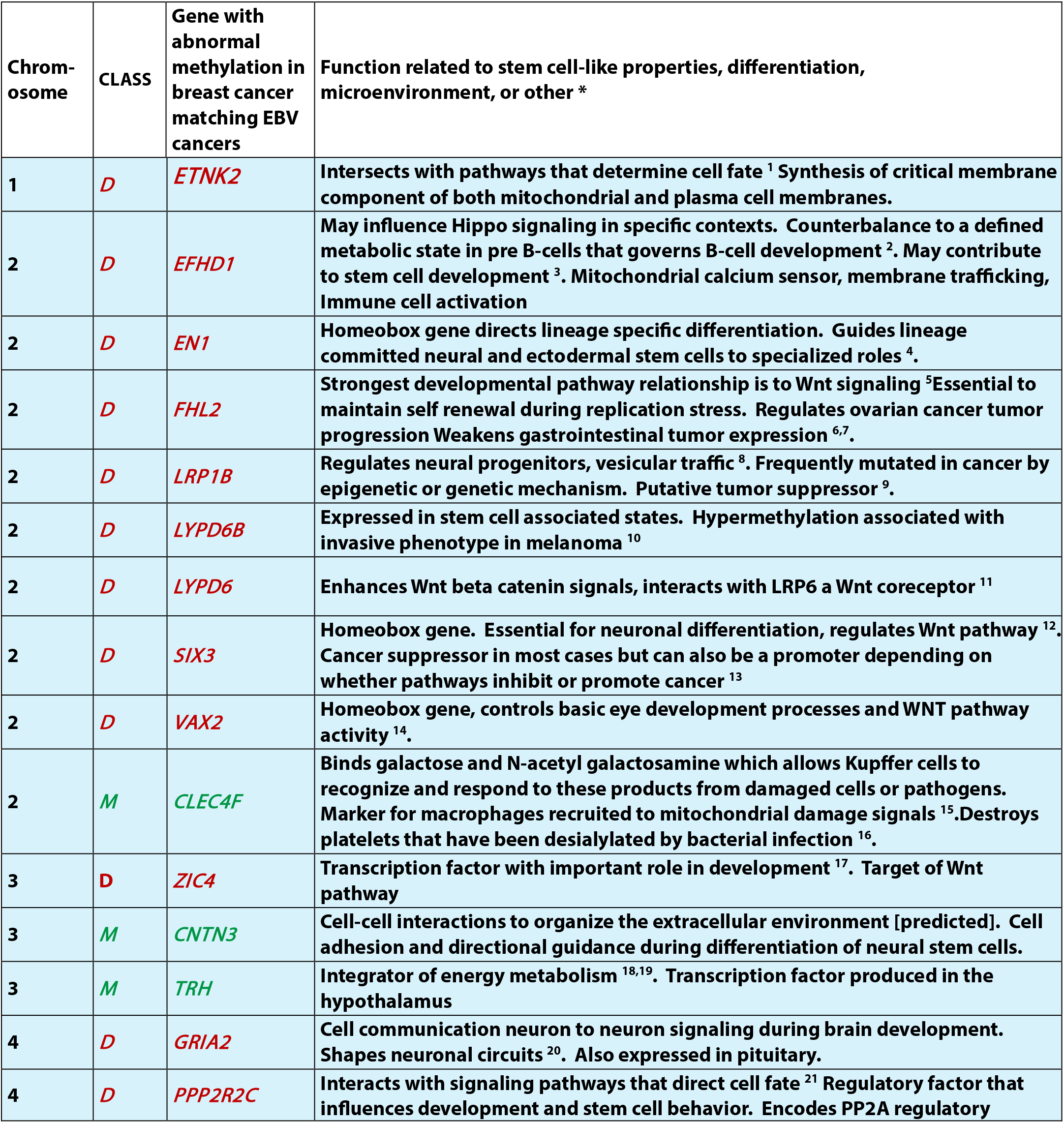
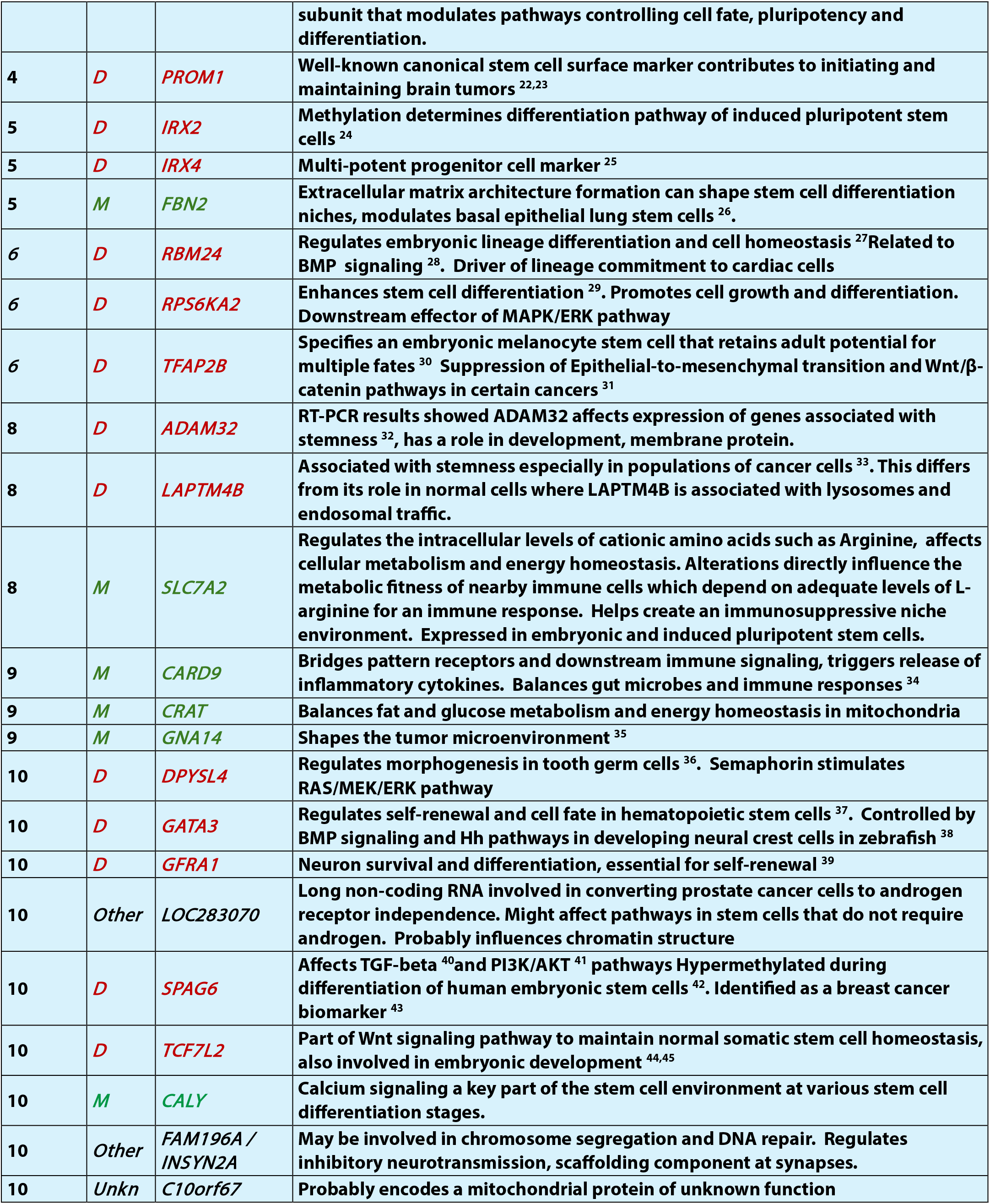
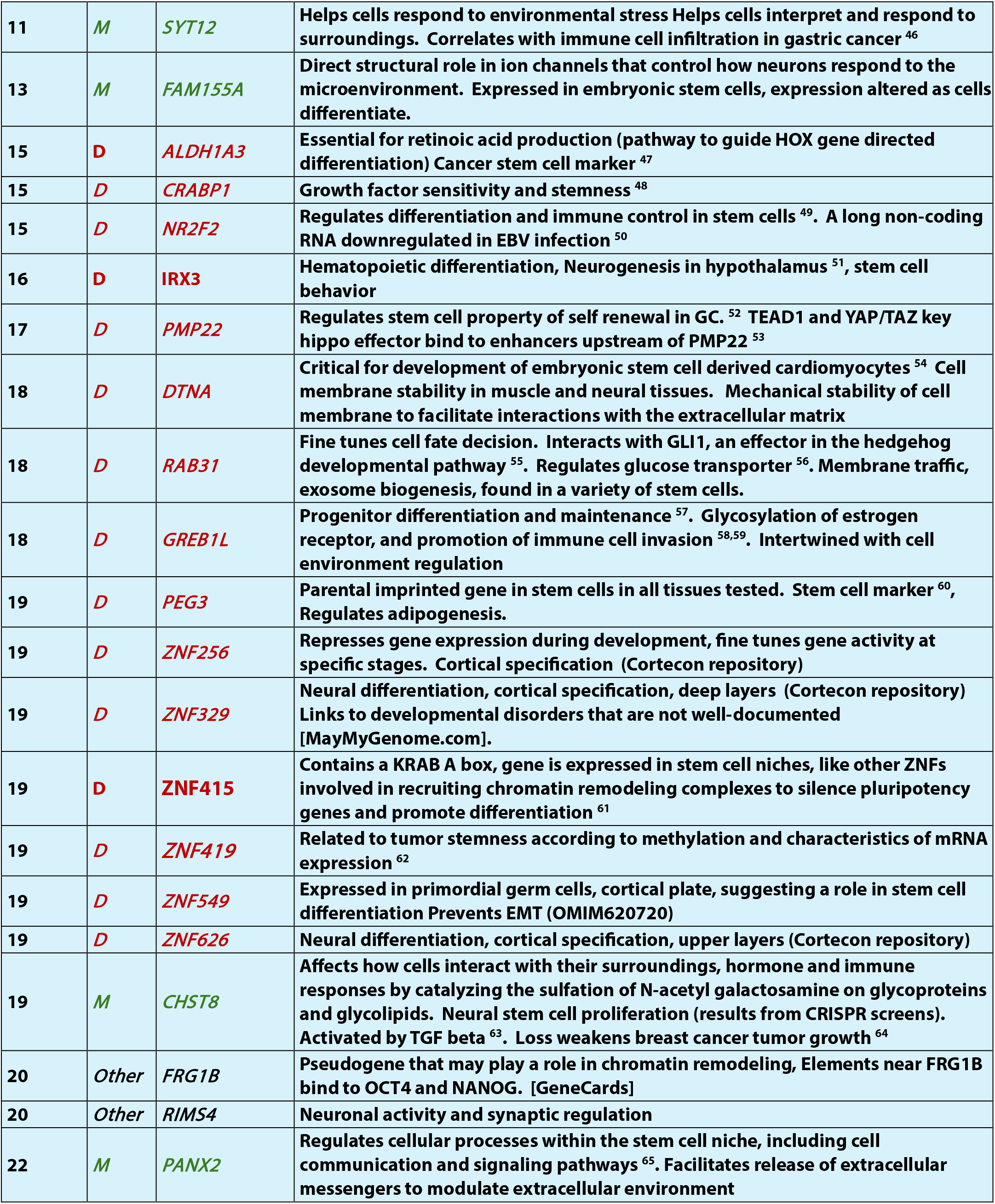
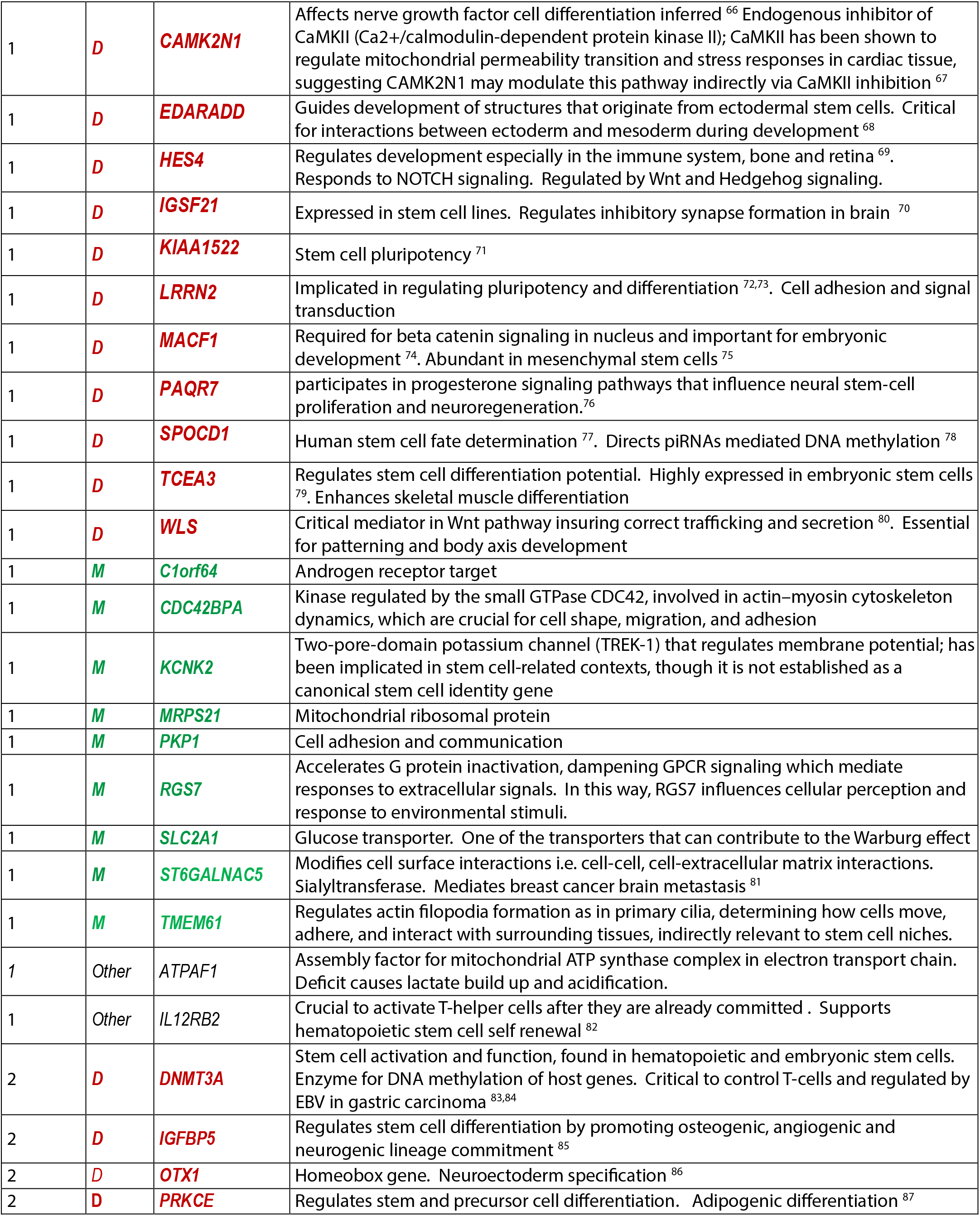
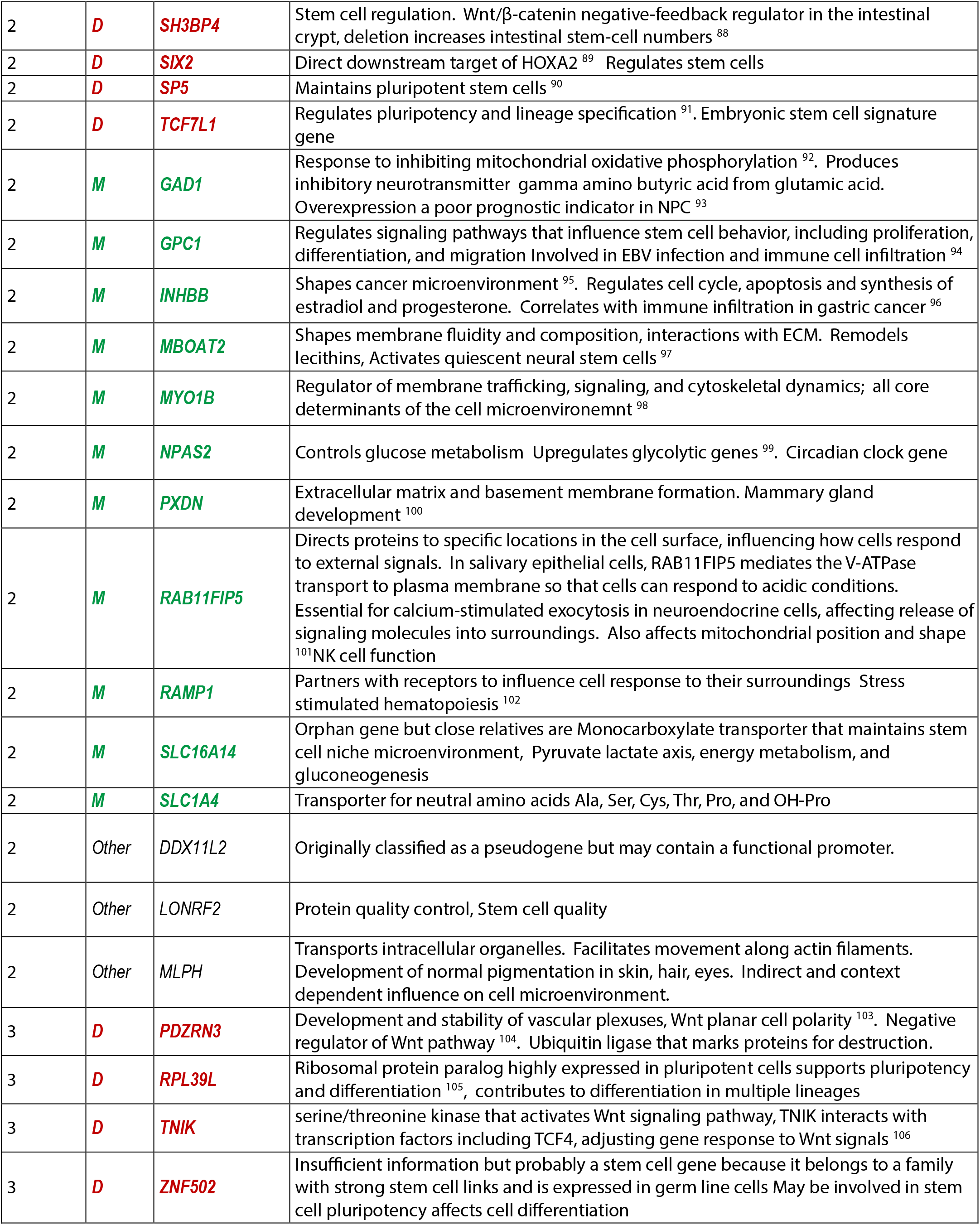
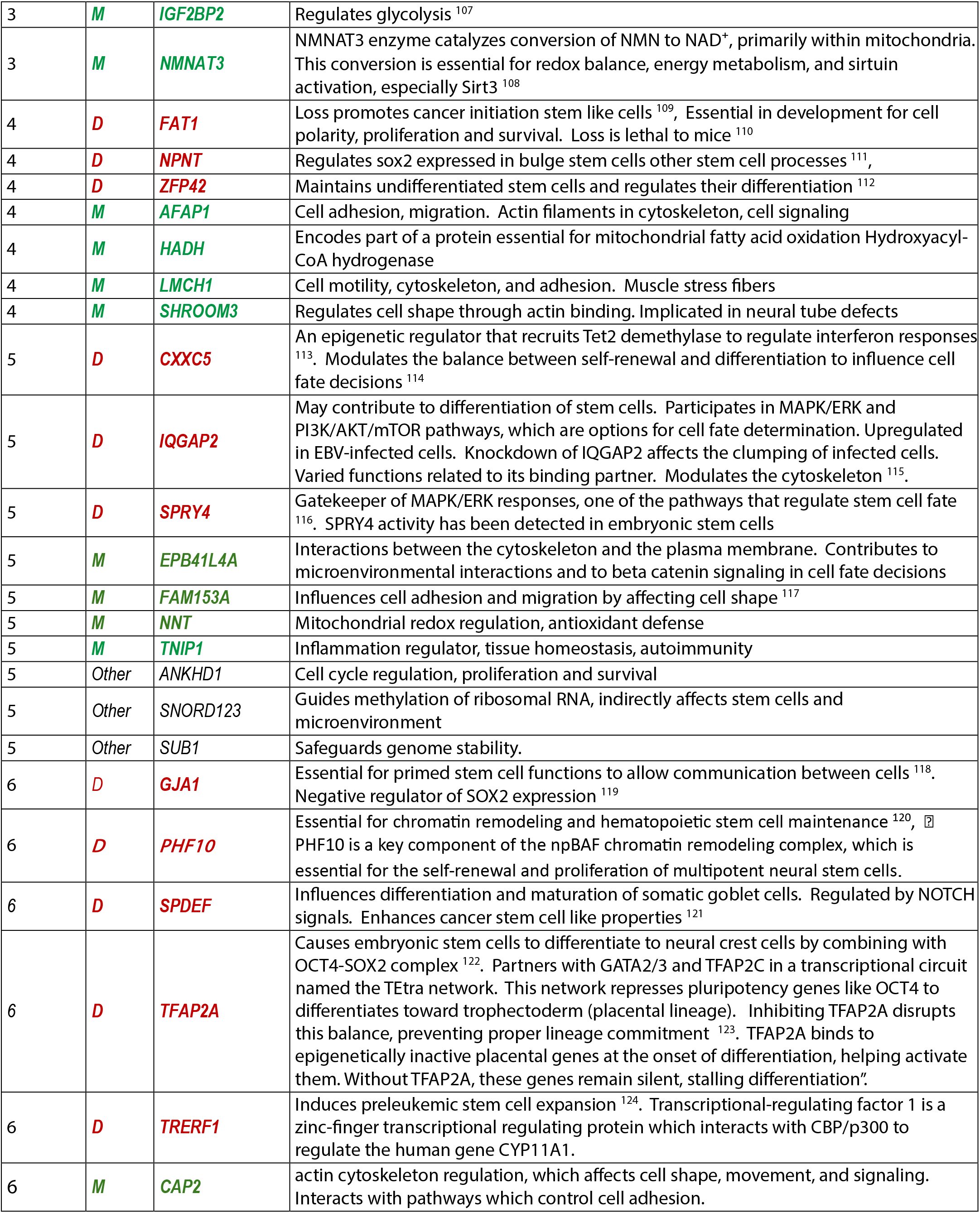
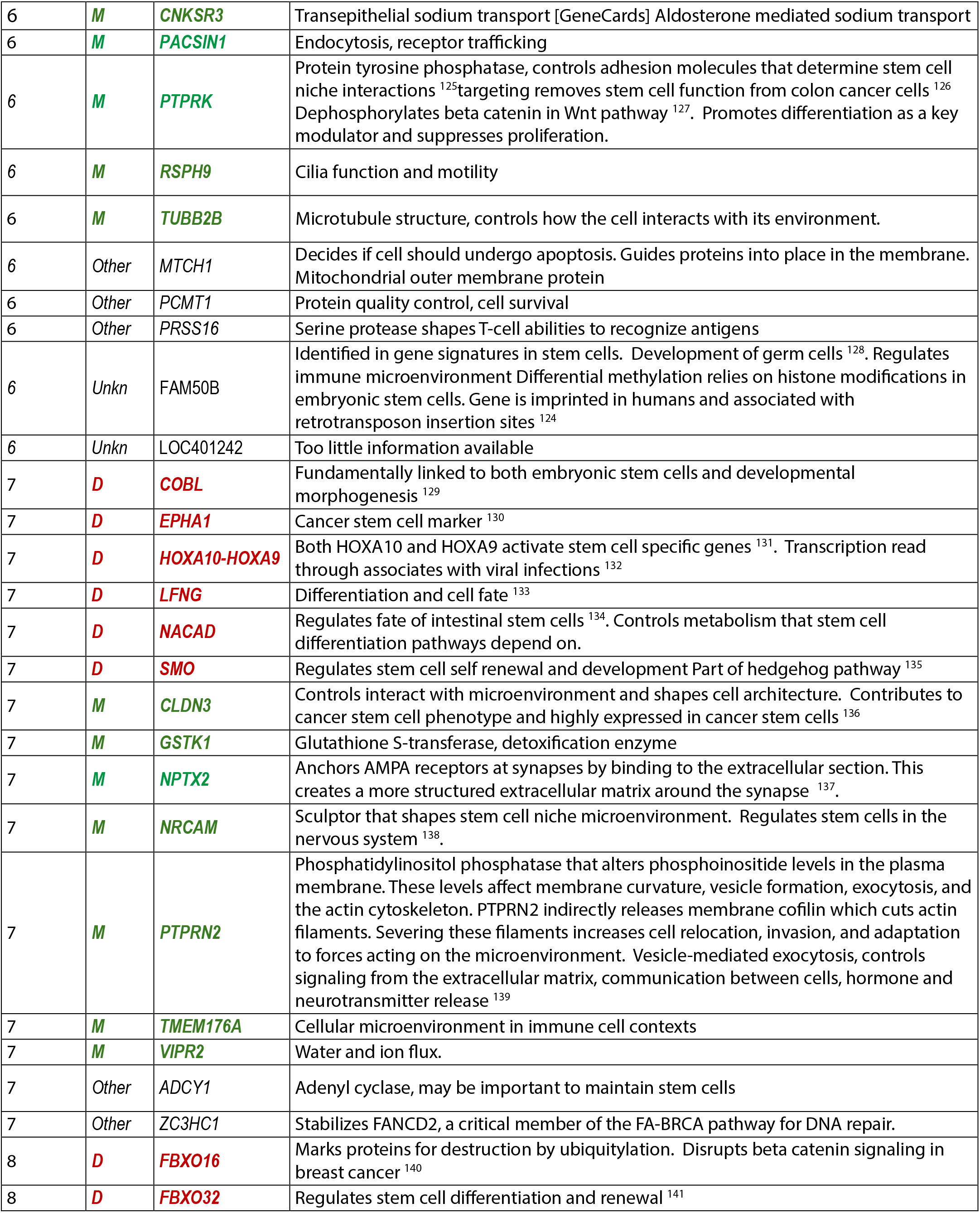
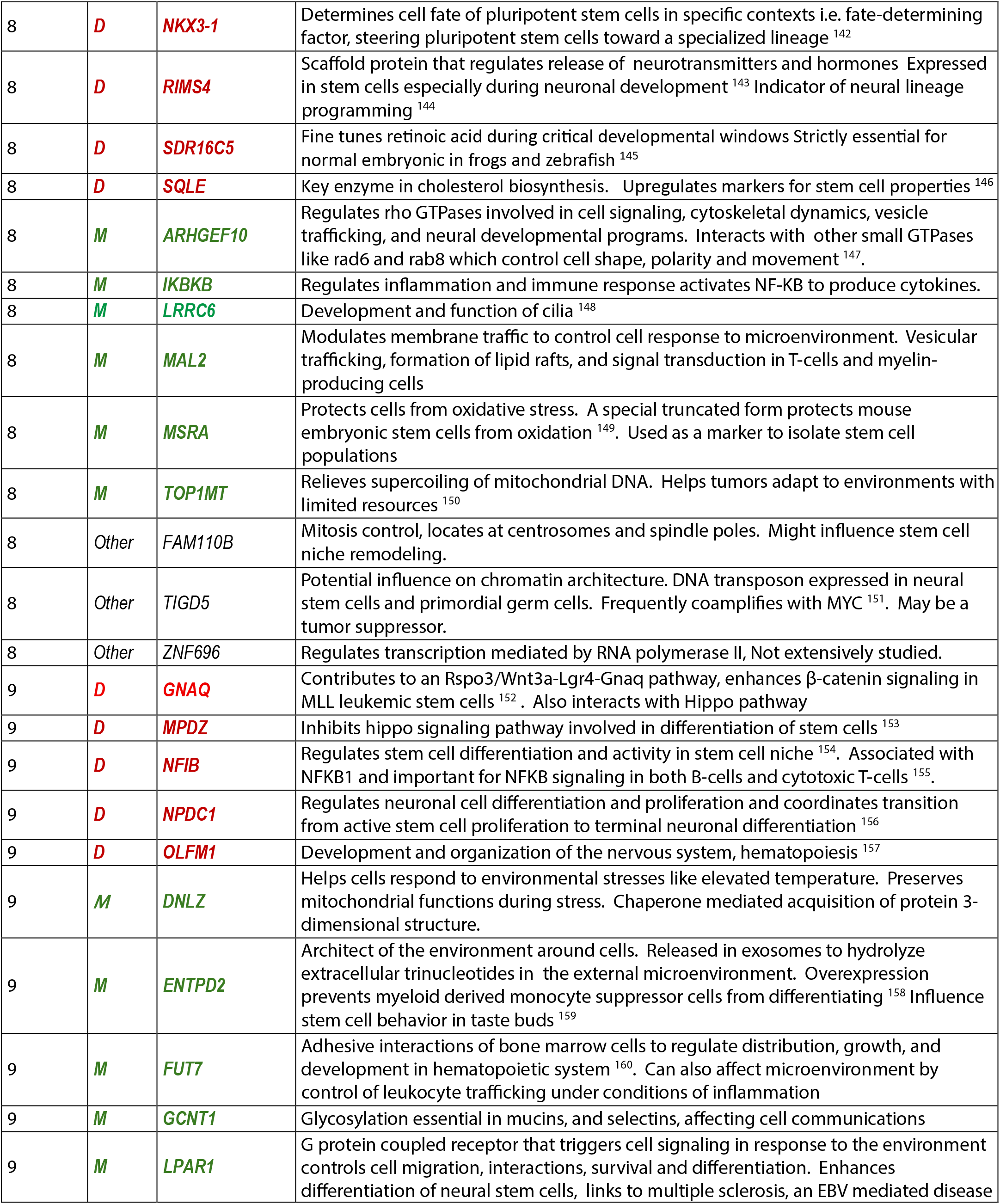
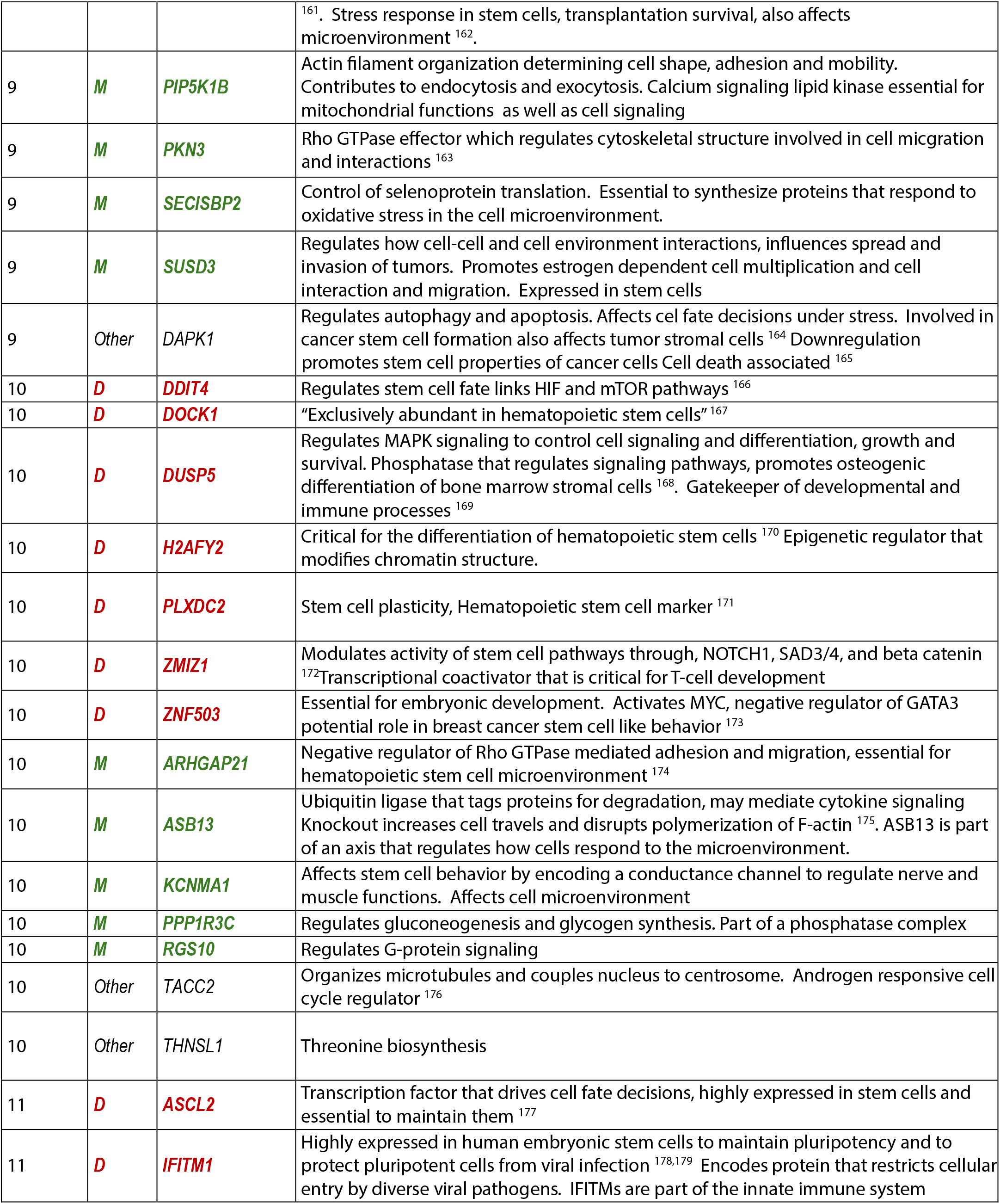
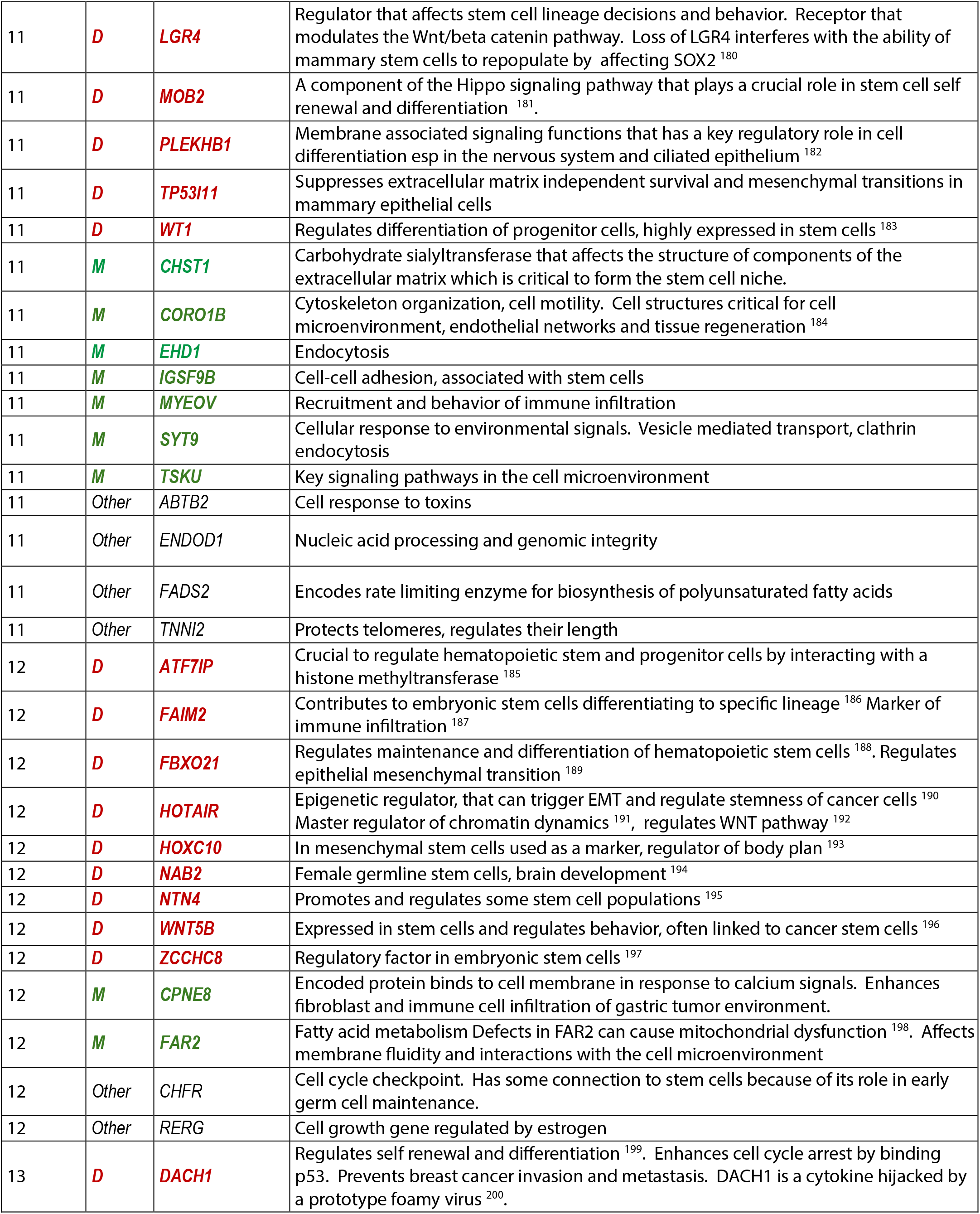
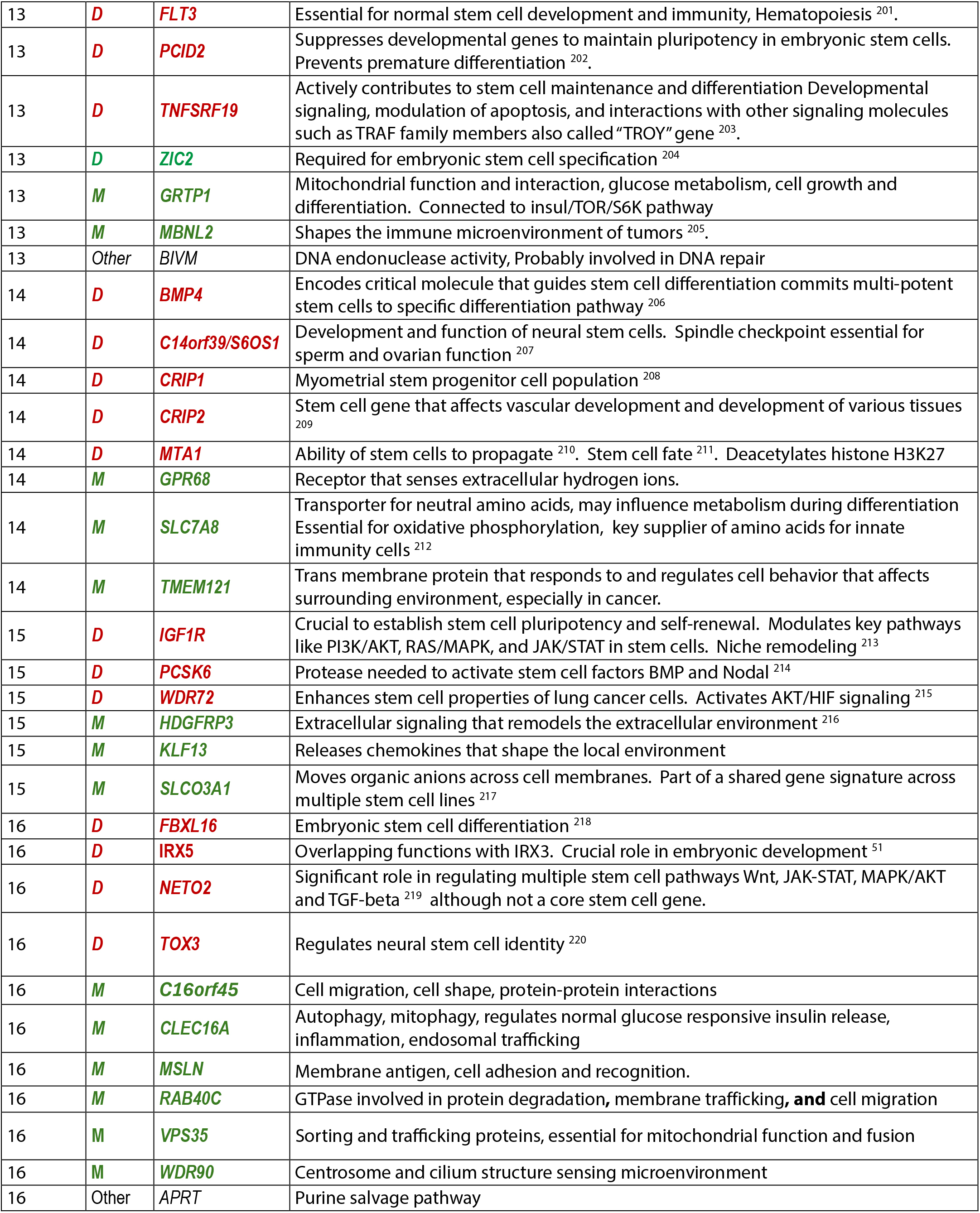
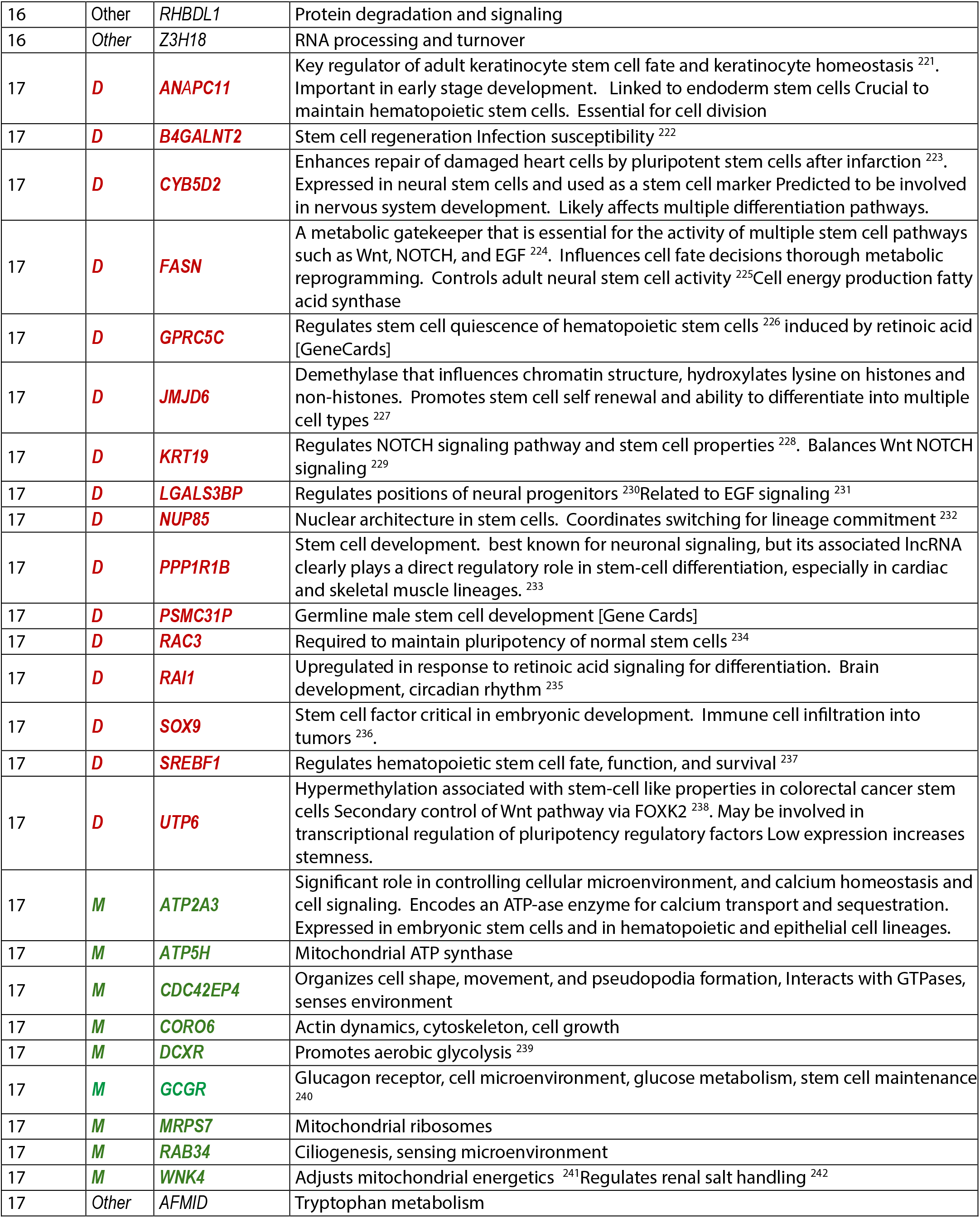
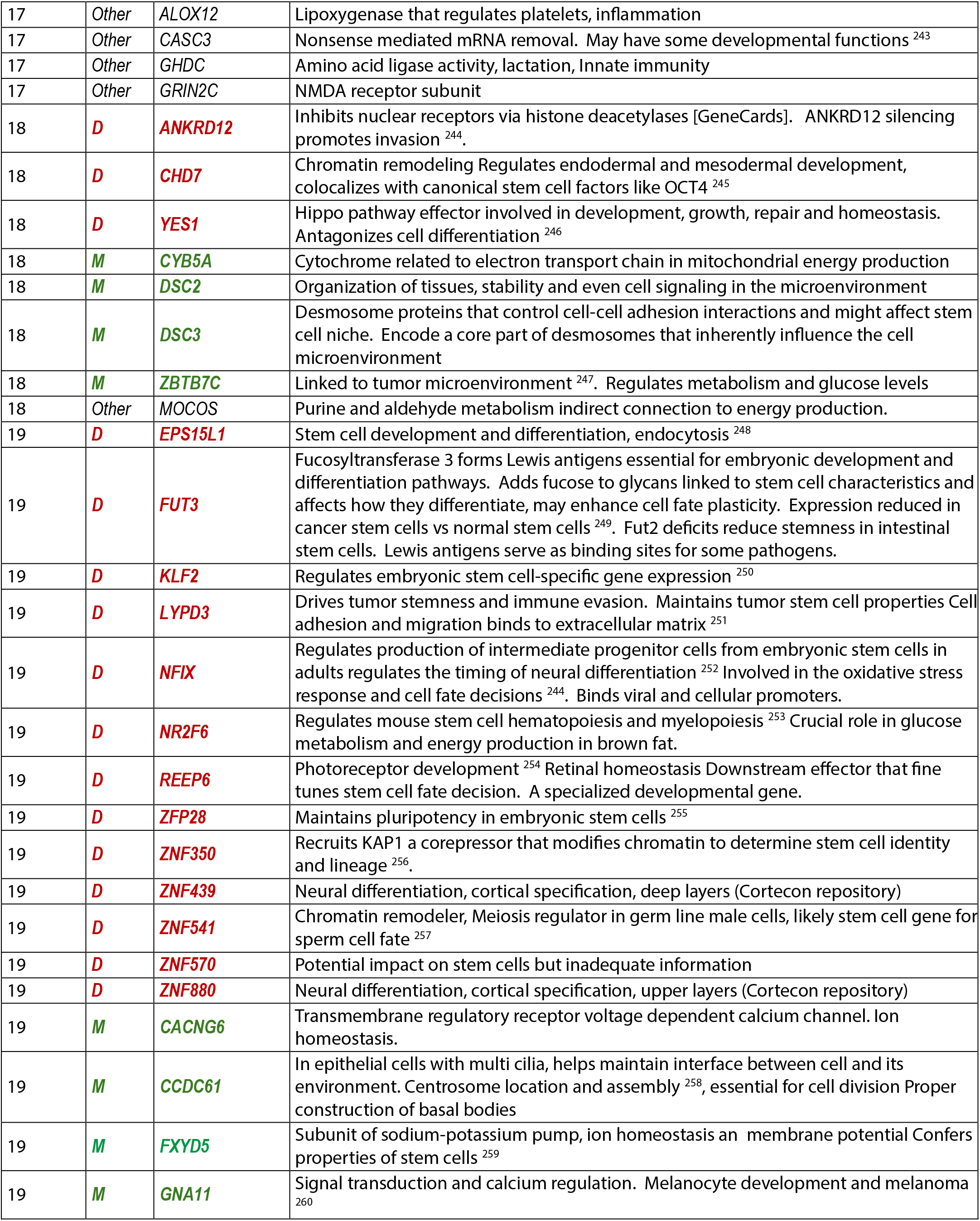
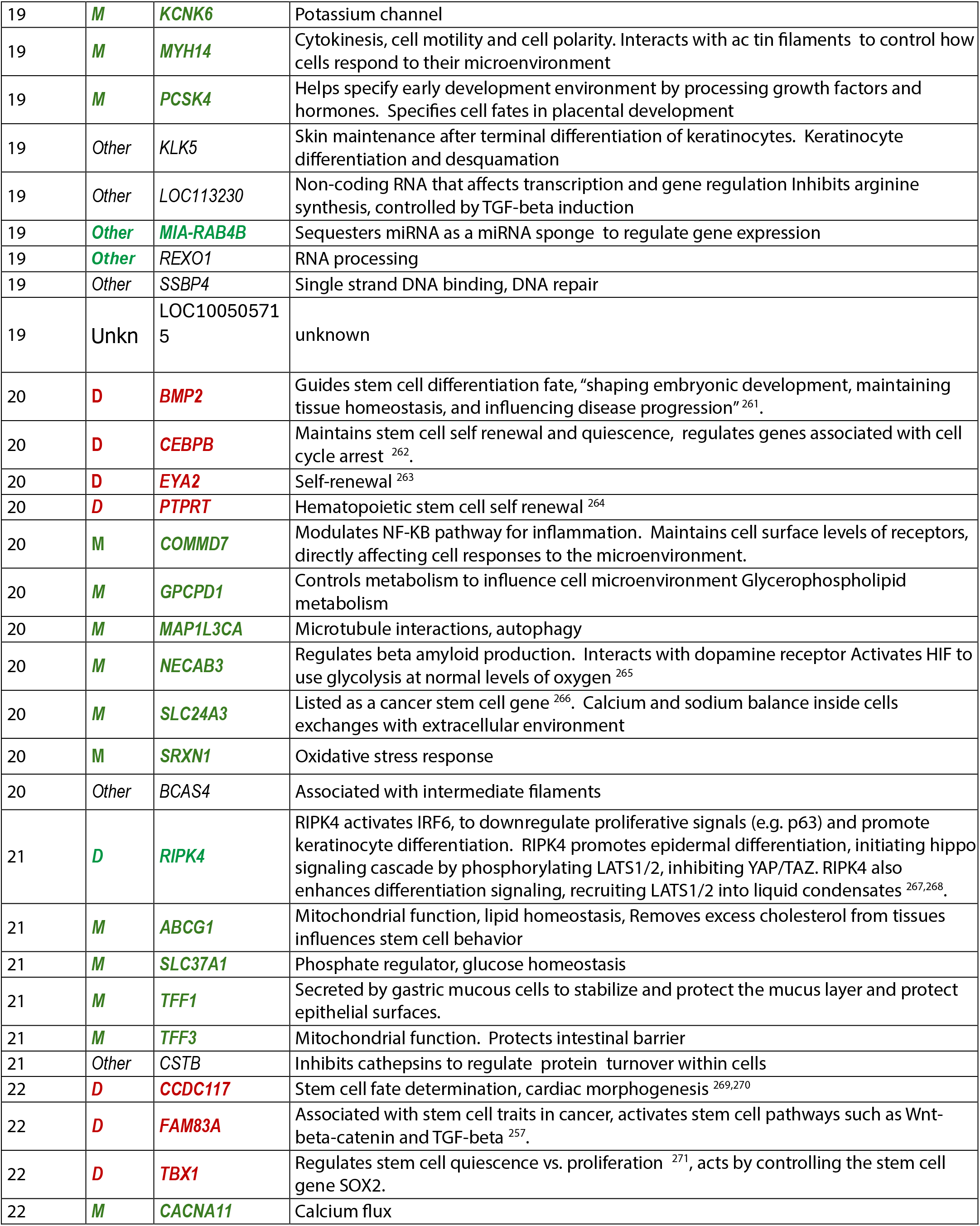
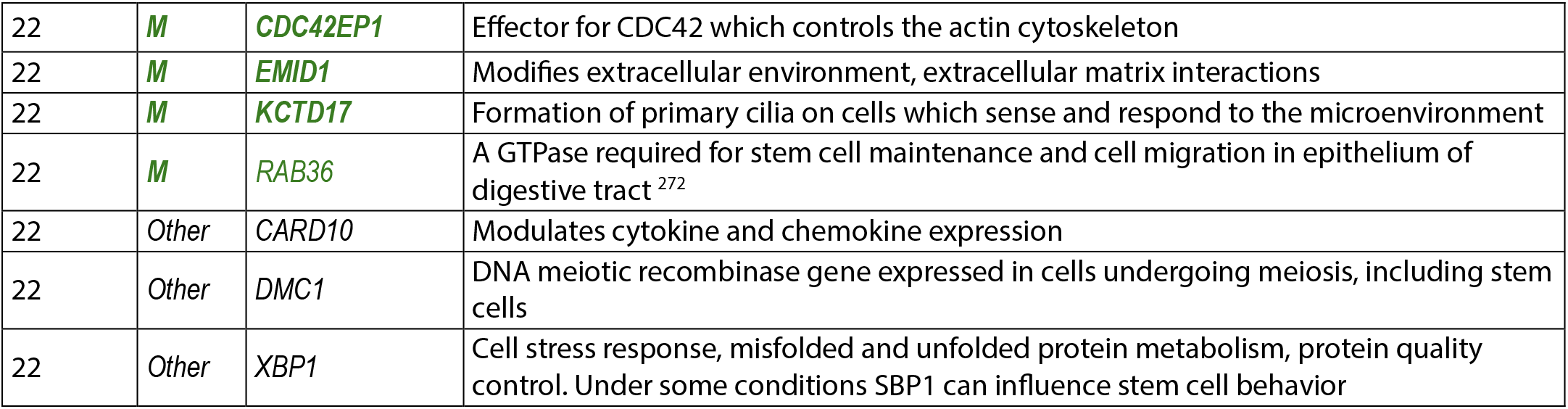
Functions at genes overlapping shared methylation loci in EBV-cancers (NPC and BL) and breast cancers. D= Genes linked to differentiation, development, or stem-like features. M=microenvironment, Other=not directly related to stem cell-like features or microenvironment. Shaded cells indicate methylation loci shared by breast cancer with both NPC and BL.

**\*Methods of gene function annotation**

Functional annotations for each gene were compiled through a structured literature review process by search for relationships to differentiation and cell microenvironment in any context. For each gene, the priority was peer-reviewed primary research articles over reviews or database summaries where possible, and selected the citation(s) that most directly and specifically supported the stated function.

Information from large databases such as NCBI is not specifically referenced. Annotations were limited to functions demonstrated by direct experimental evidence for the gene or protein in question; functional relationship was sometimes inferred indirectly (e.g., through a binding partner, upstream regulator, or downstream effector rather than the gene product itself ), this distinction was explicitly noted in the annotation text rather than presented as an established direct function.

**Extended Data Table 2.** PANTHER enrichment analysis finds potential links among cancers and developmental processes.

| PAN-GO biological process | Homo sapiens - REFLIST (20580 genes) | Number of genes (out of 57) that fit into GO term | Expected by random chance | Fold Enrichment | raw P-value | FDR |
| --- | --- | --- | --- | --- | --- | --- |
| **regulation of metabolic process (GO:0019222)** | 2815 | 20 | 7.8 | 2.57 | 3.67E-05 | 1.49E-02 |
| **regulation of macromolecule metabolic process (GO:0060255)** | 2716 | 20 | 7.52 | 2.66 | 2.17E-05 | 9.33E-03 |
| **regulation of biosynthetic process (GO:0009889)** | 2594 | 20 | 7.18 | 2.78 | 1.09E-05 | 5.00E-03 |
| **regulation of macromolecule biosynthetic process (GO:0010556)** | 2570 | 20 | 7.12 | 2.81 | 9.47E-06 | 4.66E-03 |
| **regulation of gene expression (GO:0010468)** | 2540 | 20 | 7.03 | 2.84 | 7.92E-06 | 4.55E-03 |
| **regulation of primary metabolic process (GO:0080090)** | 2434 | 20 | 6.74 | 2.97 | 4.12E-06 | 2.58E-03 |
| **regulation of nucleobase-containing compound metabolic process (GO:0019219)** | 2192 | 20 | 6.07 | 3.29 | 7.96E-07 | 7.83E-04 |
| **developmental process (GO:0032502)** | 1696 | 16 | 4.7 | 3.41 | 9.41E-06 | 4.99E-03 |
| **regulation of RNA metabolic process (GO:0051252)** | 2118 | 20 | 5.87 | 3.41 | 4.59E-07 | 6.32E-04 |
| **regulation of RNA biosynthetic process (GO:2001141)** | 1988 | 19 | 5.51 | 3.45 | 8.27E-07 | 7.12E-04 |
| **regulation of DNA-templated transcription (GO:0006355)** | 1980 | 19 | 5.48 | 3.46 | 7.78E-07 | 8.93E-04 |
| **multicellular organismal process (GO:0032501)** | 1771 | 17 | 4.91 | 3.47 | 3.58E-06 | 2.47E-03 |
| **anatomical structure development (GO:0048856)** | 1484 | 16 | 4.11 | 3.89 | 1.68E-06 | 1.29E-03 |
| **regulation of transcription by RNA polymerase II (GO:0006357)** | 1659 | 18 | 4.59 | 3.92 | 2.78E-07 | 4.78E-04 |
| **multicellular organism development (GO:0007275)** | 1073 | 16 | 2.97 | 5.38 | 2.04E-08 | 7.04E-05 |
| **system development (GO:0048731)** | 996 | 16 | 2.76 | 5.8 | 7.16E-09 | 4.93E-05 |
| **nervous system development (GO:0007399)** | 701 | 13 | 1.94 | 6.7 | 4.57E-08 | 1.05E-04 |

PAN-GO Version 2.0 Released 2025-10-12. Reference List: Homo sapiens (all genes in database).
Fisher test with FDR correction

**Extended Data Table 3.** Genes with abnormal methylation in breast cancer that share loci with EBV cancers fit into an established framework for cancer stem cells.

| Toh et al. Epigenetic Class<sup>1</sup> | Genes with shared EBV-like methylation | Cancer Stem cell function |
| --- | --- | --- |
| DNA methylation–regulated developmental transcription factors <sup>2</sup> | HOXC10, SIX2, SIX3, VAX2, TFAP2A, TFAP2B, GATA3, SPDEF, SOX9 | Stemness; lineage control; morphogen response |
| Histone modification / bivalent chromatin <sup>3,4</sup> | HOTAIR <sup>5</sup>, KLF2, TCF7L1, XBP1 | Plasticity; repression of differentiation; enhancer rewiring |
| Chromatin remodeling / architecture <sup>6</sup> | BMP2, BMP4, CRABP1, GRIA2, TOP1MT | Chromatin accessibility; RA/BMP signaling; metabolic–epigenetic coupling |
| Developmental signaling with epigenetic integration <sup>7,8</sup> | SMO, TCF7L1, SOX9 | WNT/SHH-driven chromatin remodeling |
| Differentiation-state markers under epigenetic control <sup>9</sup> | MLPH, SPAG6, SPDEF, GATA3 | Loss of lineage identity → cancer stem cell reversion |

**Extended Data Table 4.** Expression of genes in breast cancers with EBV-like methylation is unlikely to reflect B-cell lymphocyte infiltrates. Representative genes expressed in breast cancers that share methylation loci with EBV-cancers are rarely or never expressed in B-lymphocytes.

| Gene | Primary Function unrelated to B-cells (HPA dataset) | Site of Protein or RNA expression |
| --- | --- | --- |
| **ADAM32** | Metalloprotease involved in sperm maturation | Testis |
| **CNTN3** | GPI-anchored neural cell adhesion molecule involved in axon guidance | Brain, nasopharynx, Female reproductive system, heart muscle. |
| **EN1** | Homeobox transcription factor specifying midbrain-hindbrain identity | Skin |
| **GRIA2** | AMPA-type ionotropic glutamate receptor subunit mediating fast synaptic transmission | Neurons throughout the brain |
| **IRX2** | Iroquois homeobox transcription factor involved in neural tube patterning | High expression in salivary gland, seminal vesicles, and breast. Low expression in appendix and tonsils. |
| **IRX4** | Iroquois homeobox factor specifying ventricular chamber identity | Esophagus, Heart muscle, Skin, Vagina (RNA) |
| **SIX3** | Homeobox transcription factor regulating anterior brain and eye development | Forebrain and retina (RNA) |
| **SPAG6** | Structural component of the sperm flagella axoneme | Brain, Nasopharynx, Fallopian tube |
| **SYT12** | Synaptotagmin family member involved in synaptic vesicle fusion regulation | Brain, testis |
| **TRH** | Peptide hormone precursor that stimulates TSH and prolactin release | Brain (hypothalamus), endometrium, cervix |
| **VAX2** | Homeobox transcription factor regulating cell fate specification | Brain, retina |
| **ZIC4** | Zinc finger transcription factor regulating cerebellar development | Cerebellum |

**Extended Data Table 5.**
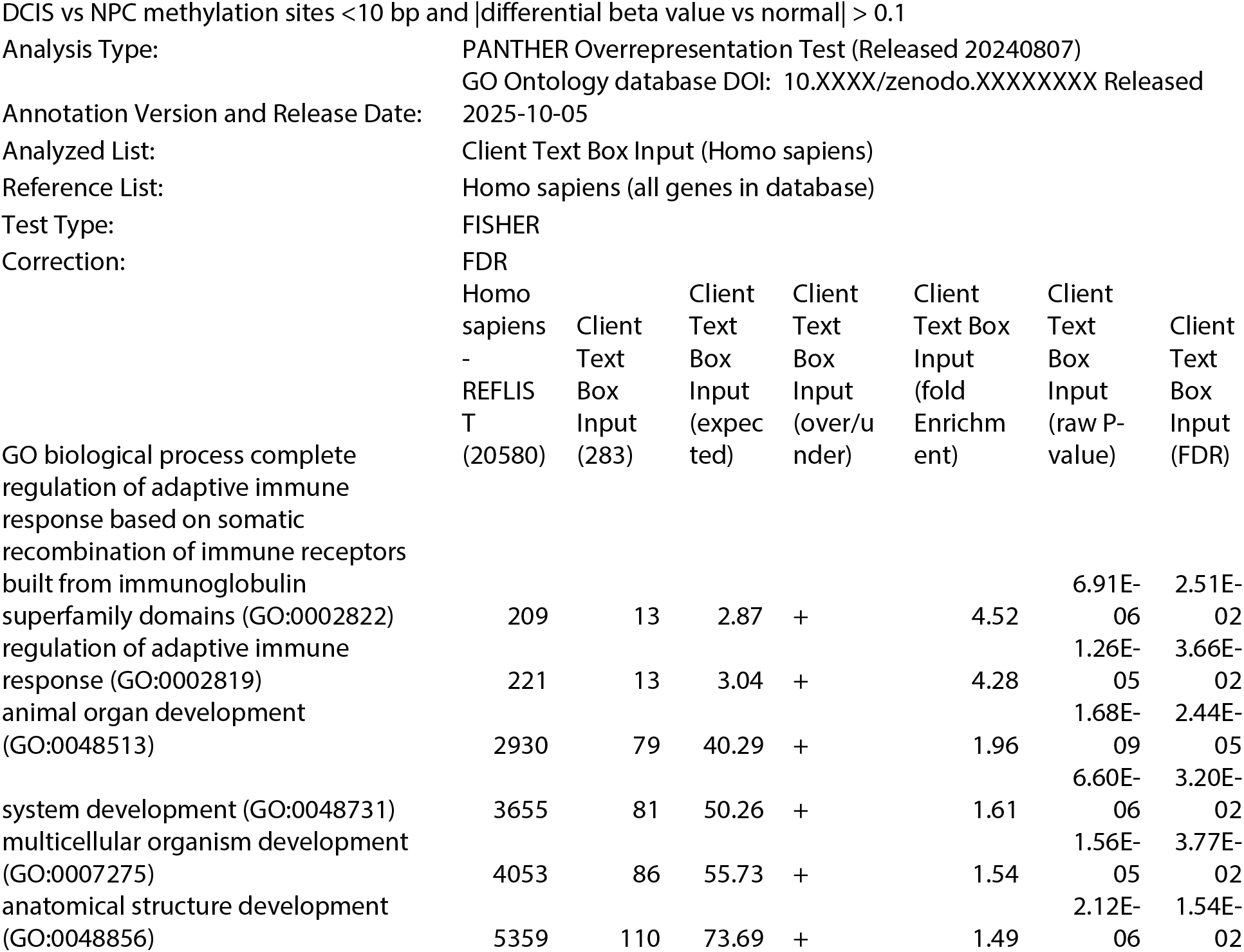
Abnormal methylation in DCIS vs NPC: PANTHER overrepresentation analysis.

**Extended Data Table 6.** Functions of genes methylated in non-malignant oral keratinocytes that are not methylated in the breast cancer cohort.

| Chromosome | Gene | Class | Relevant Encoded Function |
| --- | --- | --- | --- |
| 2 | *CYP1B1* | Microenvironment | Estradiol metabolism [1] |
| 2 | *CYP27A1* | Other | Cholesterol metabolism, bile acid synthesis [2] |
| 2 | *EMX1* | Microenvironment | Stem cell niche maintenance. Loss reduces stem cell self renewal and proliferation [3] |
| 2 | *GPR39* | Microenvironment | Disrupts tight junctions<br>microenvironment remodeling [4] |
| 2 | *MCEE* | Other | Metabolic enzyme essential for energy production in mitochondria. Metabolic rewiring |
| 4 | *CHRNA9* | Microenvironment | Ion channel receptor for acetyl choline expressed in stem cells |
| 5 | *EGFLAM* | Microenvironment | Extracellular matrix organization [5] |
| 5 | *STC2* | Microenvironment | Calcium phosphate homeostasis [6] |
| 5 | *ZNF354A* | Other | Transcription regulator cancer biomarker |
| 6 | *MB21D1* | Other | Antiviral defense, DNA damage sensor |
| 7 | *KCNH2* | Microenvironment | Voltage gated potassium channel. Activity strongly affects microenvironmental conditions. |
| 8 | *HEY1* | Stem cell | Downstream target of Notch signaling pathway, regulates self renewal and differentiation [7] |
| 8 | *NAT2* | Other | Drug metabolism and detoxification [8] |
| 8 | *ZNF1* | Other | Transcription regulation |
| 9 | *GLDC* | Microenvironment | Glycine catabolism, redox balance suppresses MHC-1 expression [9] |
| 9 | *SLC1A1* | Microenvironment | Glutamate transporter essential for neural stem cell self-renewal [10] |
| 9 | *TPRN* | Other | Inner ear function, cell structure and function |
| 10 | *GOT1* | Microenvironment | Glutamate level regulation, redox balance [11] |
| 10 | *RNLS* | Microenvironment | Regulates cardiac function, blood pressure. Cytokine like molecule that also regulates immune microenvironment [12]. |
| 10 | *UTF1* | Stem cell | Maintains pluripotency in embryonic stem cells [13] |
| 11 | *DAGLA* | Microenvironment | Affects lipid network and hormone and immune microenvironment in the CNS [14] |
| 11 | *MAP4K2* | Stem cell | Member of a pathway that affects cell fate of pluripotent stem cells. Links stress and Hippo signaling to cell fate decisions [15, 16] |
| 11 | *MUC6* | Microenvironment | Forms gelatinous barrier to protect epithelial cells, alters cell signaling, modifies ECM and immunity [17] |
| 14 | *GPX2* | Microenvironment | Redox balance and detoxification [18] |
| 15 | *ANPEP* | Microenvironment | Shapes immune cell microenvironment. Regulates microglial activation and neuroinflammation [19] Endocytosis, amino acid and peptide transport. Used as a stem cell marker |
| 16 | *MT3* | Microenvironment | Detoxification, cell response to stress, zinc homeostasis [20]. May be involved in deciding cell fate. |
| 16 | *POLR3K* | Other | Transcription, Subunit in RNA polymerase III. |
| 17 | *CLDN7* | Microenvironment | Encodes major tight junction proteins. Essential regulator for stem cell self renewal and differentiation [21] |
| 17 | *RAC3* | Stem cell | Required to maintain pluripotency of normal stem cells [22] |
| 17 | *RTN4RL1* | Microenvironment | Signaling cascades that impact actin cytoskeleton. May have stem cell like properties in nervous system but too little information available [HPA] |
| 19 | *EPOR* | Stem cell | Stem cell differentiation especially in erythrocyte and neural progenitor cells [23]. |
| 19 | *GAMT* | Other | Creatine synthesis, neurologic function [24], muscle function, energy metabolism |
| 19 | *ZNF576* | Other | Transcription regulation |
| 22 | *A4GALT* | Microenvironment | Production of a glycolipid for plasma cell membranes. Membrane composition and receptor availability for cell interactions [25]. |
| 22 | *MAPK8IP2* | Stem cell | Affects cell fate decisions in pluripotent neural progenitor cells, probably affects JNK signal pathway [26] |
| 22 | *RASD2* | Stem cell | Acts as a brake on stem cell differentiation pathways [27] |
| 22 | *SOX10* | Stem cell | Stem cell regulator and marker, mammary stem cells [28] |

**Extended Data Table 7.** Two aberrantly methylated genes shared with NPC in MS non-pathologic tissue that affect cell differentiation.

| Gene group in non pathologic MS tissue at or near NPC methylated loci | Relation to differentiation, stem cells |
| --- | --- |
| **HOXD3** | Multiple HOX genes are known EBV-responsive loci in NPC, including HOXA2 <sup>1</sup>, HOXA10 <sup>2</sup>, HOXA13 <sup>3</sup>, and HOXC8 <sup>4</sup>, which undergo EBV-associated methylation. HOXD3 links breast cancer and the CNS through a shared program of cellular plasticity: it drives de-differentiation and stemness in breast cancer cells <sup>5</sup>. HOXD3 in the CNS can reprogram endothelial cells toward angiogenic, permissive states <sup>6</sup>. |
| **NDRG4** | NDRG4 actively turns on genes essential for cell differentiation <sup>7</sup>, stabilizes neural lineage identity and restrains epithelial plasticity, expressed in brain <sup>8</sup>. NDRG4 is abnormally methylated in gastric cancer (which can be associated with EBV) <sup>9</sup>. NDRG1 is a core regulatory gene for NPC <sup>10</sup> but NDRG4 has not been studied in NPC. |

**Extended Data Table 8.** Differential NPC methylation that matches abnormal MS glial cell methylation in NAWM affects cell differentiation /development.

| **Chr** | **Genes with abnormal methylation sites in glial cells that match a site in NPC within <=10 bp** | **Relationship to cell differentiation, lineage determination** |
| --- | --- | --- |
| **1** | **PRDM16** | **Physiologic regulator of hematopoietic stem cells <sup>1</sup>. Promotes progression of neural progenitors <sup>2</sup>Controls a two direction cell fate switch between skeletal myoblasts and brown fat cells <sup>3</sup>** |
| **1** | **HS2ST1** | **Essential for neurodevelopment <sup>4</sup>** |
| **6** | **NKAPL** | **Controls differentiation factors and NOTCH signaling and stem cell markers in spermatogenesis <sup>5</sup>** |
| **7** | **HOXA3** | **Guides lineage commitment and differentiation in multiple systems and is essential to develop the third pharyngeal pouch <sup>6</sup>. Controls development of pharyngeal organs <sup>7</sup>** |
| **7** | **HOXA6** | **Potentiates proliferation of hematopoietic cells and extends self renewal <sup>8</sup>** |
| **7** | **NUPR2** | **Homeodomain transcription factor that mediates dopaminergic neuronal differentiation <sup>9</sup>** |
| **17** | **SP6** | **A signal that controls the activation, proliferation and adhesion of T-cells <sup>10</sup>** |
| **17** | **TBX2** | **Governs commitment to surface epithelium development by mediating epigenetic remodeling <sup>11</sup>** |
| **17** | **UTS2R** | **A molecular marker for intervertebral disc progenitor cells <sup>12</sup>** |
| **20** | **MATN4** | **Extracellular matrix adaptor protein that acts as a negative regulator of hematopoietic stem cell stress-induced proliferation and serves as a dynamic transcriptional marker during lineage progression and differentiation in glial progenitor populations like oligodendrocyte precursor cells <sup>13</sup>** |
| **20** | **RBPJL** | **Controls response of cells to Notch activation<sup>14</sup>. Essential for osteoblast differentiation <sup>15</sup>** |

**Extended Data Table 9.** Differentially methylated genes (delta beta>0.2) in whole blood from MS patients that match differentially methylated genes in NPC.

| Gene with strong evidence for differentiation role | Chromosome | Developmental / Differentiation Role | Key Supporting Citations |
| --- | --- | --- | --- |
| **RHOH** | 4p14 | Stem cell regulation | [1] |
| **PITX2** | 4q25 | Craniofacial and ocular development | [2, 3] |
| **SCGB3A1** | 5q31.1 | Promotes muscle fiber differentiation | [4] |
| **DOK3** | 5q35.1 | Promotes plasma cell differentiation | [5] |
| **HOXA9** | 7p15.2 | Essential for hematopoietic stem cell self renewal and emergence | [6, 7] |
| **HOXA10** | 7p15.2 | Positional identity in stem cells | [8-10] |
| **FEZF1** | 7q31.2 | Essential for neural differentiation | [11] |
| **LRP5** | 11q13.2 | Coreceptor for the major developmental pathway Wnt/beta catenin | [12] |
| **TNFSF12** | 17p13.1 | Shapes cell fate decisions | [13] |
| **ZZEF1** | 17q12 | Histone reader. Coregulator of KLFs, which are central regulators of pluripotency, differentiation and development | [14] |
| **FGF11** | 17q22 | Neuronal development and differentiation. Regulate stem cell pluripotency | [15, 16] |

**Extended data Table 10.** Functions of genes with the same abnormal methylation loci in non pathologic MS brain tissue and breast cancer Genes are grouped under functional theme with the strongest supporting documentation.

| Gene | Chromosome | Documented role in differentiation | Reference |
| --- | --- | --- | --- |
| **RUNX3** | 1 | Runt-domain transcription factor controlling T-cell lineage differentiation (CD4/CD8 lineage choice) and epithelial/neuronal differentiation programs | 1,2 |
| **FOXO6** | 1 | Forkhead-family transcription factor implicated in neuronal differentiation and metabolic regulation; family members regulate stem cell quiescence/self-renewal | 3 |
| **MSX1** | 4 | Homeobox transcription factor that maintains progenitor/undifferentiated states and regulates craniofacial, tooth, and myogenic differentiation decisions | 4 |
| **SPI1 (PU.1)** | 11 | Master regulator of hematopoietic stem/progenitor cell fate; required for myeloid and B-lymphoid lineage development from multipotent progenitors | 5,6 |
| **SOX8** | 16 | SoxE-family transcription factor involved in glial, Sertoli cell, and osteoblast differentiation from progenitor pools; functionally redundant with SOX9/SOX10 | 7,8 |

| Gene | Chromosome | Documented role | Reference |
| --- | --- | --- | --- |
| **HHAT** | 1 | Hedgehog acyltransferase; palmitoylates Sonic/Indian Hedgehog ligands, a modification required for their long-range signaling activity that drives progenitor differentiation in many tissues | 9 |
| **MAML3** | 4 | Member of the Mastermind-like family of Notch co-activators; required for transcriptional output of Notch signaling, a pathway that governs binary differentiation decisions in numerous stem/progenitor systems | 10 |

3. Chromatin and Epigenetic Regulators Gating Differentiation Programs
| Gene | Chromosome | Documented role | Reference |
| --- | --- | --- | --- |
| **PHC2** | 1 | Polycomb-group protein (Polyhomeotic homolog 2), component of PRC1 complex involved in chromatin compaction and repression of developmental genes | 11,12 |
| **HDAC4** | 2 | Histone deacetylase that represses differentiation gene programs (e.g., chondrocyte hypertrophy, muscle differentiation) until released by signal-dependent nuclear export | 13 |
| **DAXX** | 6 | Chromatin-associated protein involved in transcriptional repression and heterochromatin maintenance via PML nuclear bodies | 14 |
| **NNMT** | 11 | Regulates epigenetics by depleting methyl donor SAM acting as a metabolic methylation sink | 15 |

4. Positional Identity / Hox Network
| Gene | Chromosome | Documented role | Reference |
| --- | --- | --- | --- |
| **HOXD3** | 2 | HOX-cluster homeobox gene specifying positional identity along the body axis during development; Hox genes broadly pattern differentiation of progenitors along anteroposterior axes | 16 |
| **HAGLR (Hoxd-AS1)** | 2 | Long non-coding RNA transcribed from the HOXD cluster, proposed to regulate neighboring HOXD gene expression in cis | 17 |

| Gene | Chromosome | Documented role | Reference |
| --- | --- | --- | --- |
| **CD247 (CD3ζ)** | 1 | Signaling chain of the T-cell receptor complex; essential for T-cell receptor signal transduction during T-cell development | 18 |
| **NLRP3** | 1 | Core component of the NLRP3 inflammasome; drives IL-1β processing during innate immune/monocyte differentiation and activation | 19 |
| **SP140** | 2 | Epigenetic reader. Lymphoid-lineage nuclear body protein implicated in macrophage/B-cell differentiation and chromatin organization | 20 |
| **CYTIP (Cybr)** | 2 | Regulates cytohesin-1 activity and dendritic cell migration/maturation. The mechanism may control T-cell receptor activation | 21 |
| **LRBA** | 4 | Regulates intracellular trafficking of CTLA-4 in T cells; loss causes immune dysregulation, implicating it in T-cell functional differentiation/homeostasis | 22 |
| **MARCHF1 (MARCH1)** | 4 | E3 ubiquitin ligase that downregulates MHC class II surface expression during dendritic cell maturation/differentiation | 23 |
| **CCR6** | 6 | Chemokine receptor whose expression marks and functionally supports Th17 differentiation from naive CD4+ T cells | 24 |
| **TRAF3IP2 (Act1)** | 6 | Adaptor protein required for IL-17 receptor signaling, relevant to Th17 effector differentiation and downstream inflammatory signaling | 25 |
| **MPEG1 (Perforin-2)** | 11 | Antimicrobial pore-forming protein expressed during macrophage differentiation/maturation | 26,27 |
| **LSP1** | 13 | Leukocyte-specific, actin-binding protein involved in neutrophil/leukocyte motility during immune cell maturation. Critical for dendritic and CD8 T-cell functions | 28,29 |

| Gene | Chromosome | Documented role | Reference |
| --- | --- | --- | --- |
| **PKP4 (p0071)** | 2 | Armadillo-family protein at adherens junctions/desmosomes, links cadherins to the cytoskeleton. Regulates keratinocyte differentiation via the Hippo signaling pathway. | 30 |
| **MICAL2** | 11 | Actin-disassembly factor acting downstream of semaphorin/plexin signaling, regulating cytoskeletal remodeling | 31 |
| **ARHGAP1** | 11 | Rho GTPase-activating protein regulating Rho/Cdc42 signaling and actin cytoskeleton dynamics | 32,33 |
| **MCF2L** | 13 | Dbl-family guanine nucleotide exchange factor that activates Rac1/RhoA GTPases.. Controls remodeling of cytoskeleton and cell migration | 34 |
| **BCAR1 (p130Cas)** | 16 | Scaffold/adaptor protein in integrin adhesion signaling, implicated in cell migration and originally identified in antiestrogen-resistance/breast cancer signaling studies | 35 |
| **GNAO1** | 16 | G-protein alpha subunit involved in neuronal signal transduction and neurite outgrowth | 36 |
| **CDH8** | 16 | Mediates calcium dependent cell-cell adhesion in the brain it guides axon growth and helps build neural circuits | 37 |

**1. Master Transcription Factors for Lineage Commitment**

These genes encode transcription factors with well-documented, experimentally established roles in driving stem or progenitor cells toward a specific differentiated fate.

**2. Developmental Signaling Pathways Driving Differentiation**

Hedgehog and Notch are two of the core signaling pathways that instruct stem and progenitor cells to differentiate, and both are represented in this list.

**3. Chromatin and Epigenetic Regulators Gating Differentiation Programs**

**4. Positional Identity / Hox Network**

**5. Immune Cell Receptor Signaling and Lineage-Specific Immune Factors**

A large cluster of genes points to hematopoietic/immune lineage specification and effector differentiation (e.g., naive T cells differentiating into Th17 cells, myeloid progenitors into macrophages/dendritic cells).

**5. Cytoskeletal Remodeling and Cell Adhesion (Supports Morphological Change During Differentiation)**

Differentiation frequently requires coordinated changes in cell shape, adhesion, and migration. These genes are documented cytoskeleton/adhesion regulators; their connection to differentiation is generally mechanistic/supportive rather than lineagedefining.

**Extended Data Table 11.** Abnormal NAWM methylation sites shared with breast cancer genes that affect differentiation. These genes have abnormal methylation at essentially the same positions in NAWM of MS brains ^1^ and in breast cancer ^2^. The research cited, demonstrates these genes have clear, established roles in differentiation, morphogenesis, developmental patterning, or reprogramming.

| Gene | Chromosome | Relationship | Reference |
| --- | --- | --- | --- |
| CASZ1 | 1p36.22 | Zinc-finger TF; cardiac precursor cell differentiation | 3 |
| GRHL3 | 1p36.11 | Grainyhead TF; epidermal differentiation, neural tube closure. Promotes differentiation and migration | 4 |
| AHDC1 | 1p36.11 | Early epithelial morphogenesis | 5 |
| CACNA1E | 1q25.3 | Calcium channel essential for neurodevelopment | 6 |
| ROR1 | 1p31.3 | regulates developmental morphogenesis and tissue- and organogenesis; muscle cell regeneration | 7 |
| DNAJC6 | 1p31.1 | Essential for WNT pathway functions during neuron development | 8 |
| HLX | 1q41 | Regulates early hematopoiesis; visceral organogenesis | 9,10 |
| NKAPL | 6p22.1 | Controls differentiation factors and NOTCH signaling and stem cell markers in spermatogenesis | 11 |
| CLIC5 | 6p21.1 | Promotes cell differentiation and regeneration | 12 |
| HOXA3 | 7p15.2 | Guides lineage commitment and differentiation in multiple systems and is essential to develop the third pharyngeal pouch . Controls development of pharyngeal organs | 13,14 |
| HOXA6 | 7p15.2 | Potentiates proliferation of hematopoietic cells and extends self renewal | 15 |
| STEAP1B | 7q21.13 | Metal ion homeostasis. Regulates cell differentiation | 16 |
| PTPRN2 | 7q36.3 | Consistently linked to endocrine cell differentiation | 17 |
| OR51B5 | 11p15.4 | Epidermal differentiation | 18 |
| APBB1 | 11q23.3 | Cell fate decisions | 19 |
| MIR100HG | 11q24.1 | Long non-coding RNA host gene that regulates cell differentiation, development, and stem cell programming by interacting with major signaling pathways and embedded microRNAs | 20 |
| NTM | 11q25 | Nervous system development, cell differentiation | 21 |
| CORO7-PAM16 | 16p13.3 | Essential for bone development | 22 |
| CORO7 | 16p13.3 | Key controller for Hippo pathway which controls organ size and tissue homeostasis. Activates Hippo pathway in response to cell-cell contact | 23 |
| GRIN2A | 16p13.2 | Mutations cause neurodevelopmental disorders | 24 |
| BCAR1 | 16q23.1 | Clear role in somatic cell differentiation | 25,26 |
| STAT5A | 17q11.2 | Stem cell self renewal, bias stem cells into erythroid lineage. | 27 |

**Extended Data Table 12.**
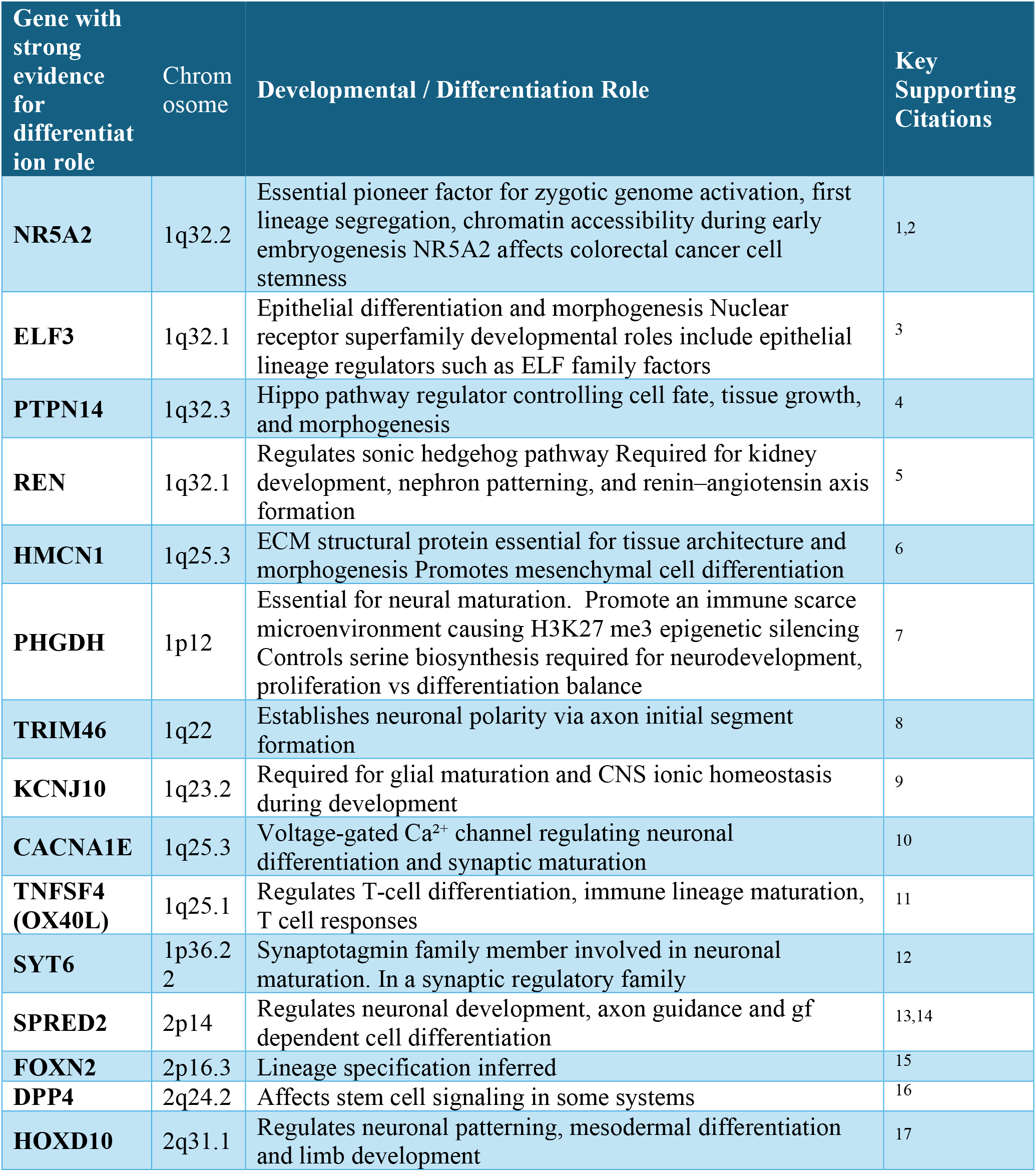
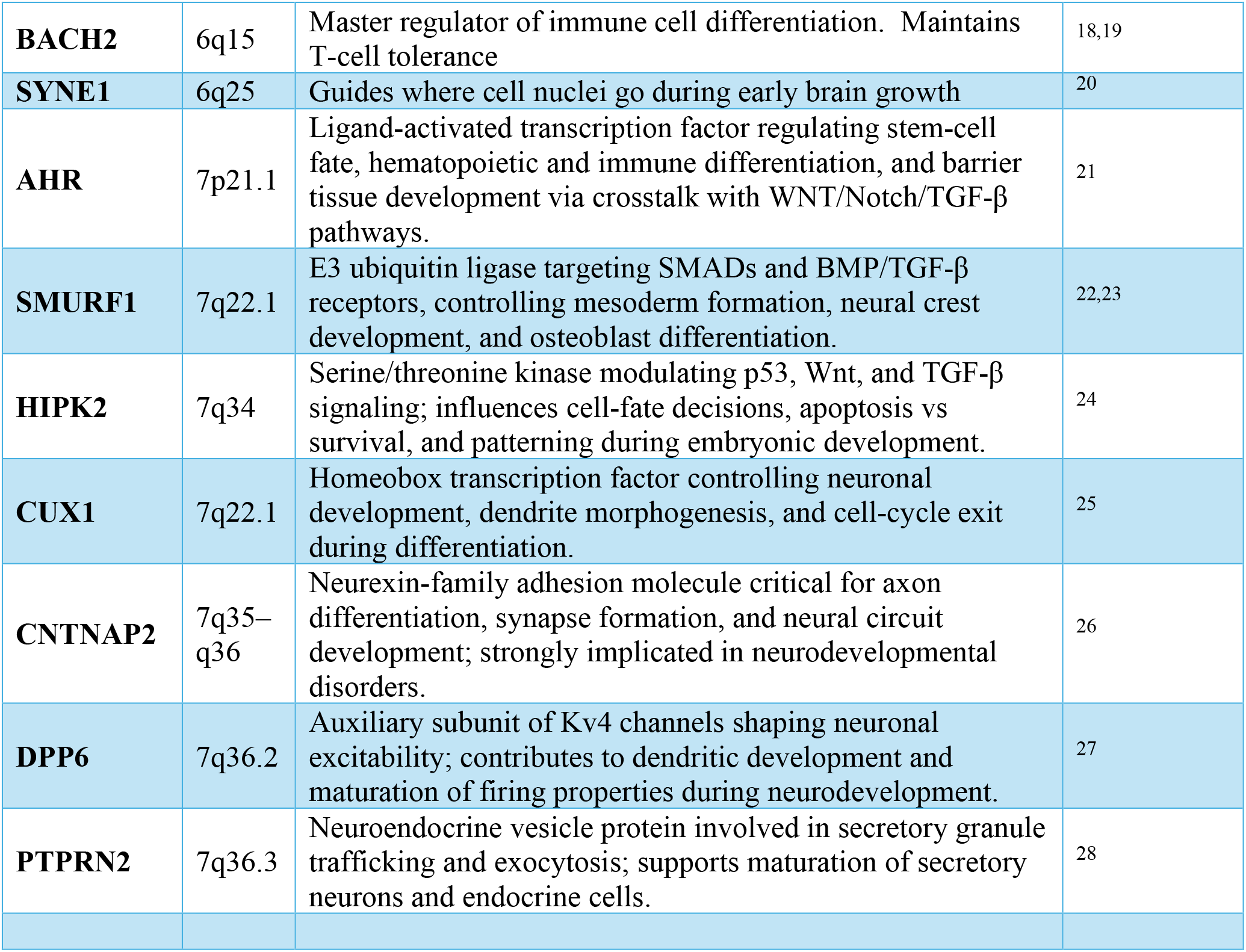
Differentially methylated genes (delta beta>0.2) in MS whole blood that match differentially methylated genes in breast cancer and have strong links to cell differentiation/development.

